# The Hidden Burden of Mortality Across the Spectrum of ICD-10 Conditions in Australia: A Multiple Cause of Death Analysis

**DOI:** 10.64898/2026.02.07.26345820

**Authors:** Hayden Farquhar

## Abstract

**Background:** Death certificates record both an underlying cause and contributing conditions, yet mortality statistics predominantly report only the underlying cause. We quantify this “hidden burden” across all ICD-10 conditions in Australian mortality data using the multiple-to-underlying ratio (MUR): total death certificate mentions divided by underlying cause deaths.

**Methods:** We analysed Australian Bureau of Statistics Causes of Death 2023 data (N = 187,268 registered deaths) to compute the ratio for all ICD-10 conditions. Three pre-registered confirmatory hypotheses tested sex differences in hypertension and mental health ratios, and geographic variation by preventability, with Holm-Bonferroni correction.

**Results:** Death certificates recorded an average of 3.5 causes per decedent, meaning the underlying cause captures only ∼29% of recorded morbidity. Of 663 conditions with ≥10 underlying cause deaths, the ratio ranged from 1.0 (external causes) to 281.1, with a median of 2.5. Among conditions with stable estimates (≥50 underlying deaths), the highest ratio was 94.3 (complications of medical care). Age explained only 10.9% of ratio variation (R^2^ = 0.109), and no top-ranked conditions were identified as primarily age-driven, suggesting the ratio ranking is robust to age confounding. However, external validation using US CDC data showed age standardisation materially changed absolute ratio values for 6 of 8 cause groups (divergence 16–34%), with the direction varying by condition rather than following a simple age-concentration pattern. Males showed consistently higher ratios, most strikingly for mental health disorders (62% higher); a counterfactual analysis estimated suicide coding rules explain only 6–15% of this sex difference. Three pre-registered hypotheses were null after correction; H1 and H3 (sex differences) were underpowered (n = 4, n = 8 pairs) with large effect sizes (r = 0.77–0.80), while H2 (geographic variation) showed a clear null.

**Conclusions:** The hidden burden of mortality in Australia is substantial and unevenly distributed, with symptom codes, mental health conditions, and hypertension most undercounted. The ratio provides a transparent framework for identifying conditions whose health impact is systematically understated by conventional mortality reporting.

## Introduction

Mortality statistics are the foundation of public health surveillance, resource allocation, and health system performance measurement. In Australia, as in most countries that follow World Health Organization (WHO) conventions, published mortality data predominantly report the *underlying cause of death*, defined as “the disease or injury which initiated the train of morbid events leading directly to death” [1]. This single-cause approach, while essential for international comparability, may systematically undercount the contribution of chronic conditions that frequently participate in, but do not initiate, the causal chain of death.

Death certificates in Australia, as in other countries using the WHO International Form of Medical Certificate of Cause of Death, can record multiple conditions [1]. Part I captures the causal sequence from immediate cause to underlying cause, while Part II records other significant conditions contributing to death. The *multiple causes of death* (MCOD) framework considers all conditions mentioned on the certificate, providing a more complete picture of the morbidity burden at the time of death.

Modern death certificates typically record several conditions per decedent. International studies report averages of 3–4 causes per certificate: 3.4 in Australia [2], with similar figures in Europe [3] and the United States [4]. This means the underlying cause (the single condition selected from each certificate) represents less than one-third of the recorded morbidity at death, measured by share of mentions. This is not to equate mentions with causal weight; the underlying cause has a privileged causal role by definition. But the remaining conditions, though judged by certifying physicians to have contributed to death, are invisible in standard mortality tabulations.

The discrepancy between underlying cause and multiple cause counts has been examined across diverse settings. Foundational work in the United States demonstrated that the choice of underlying cause substantially altered the apparent ranking of disease burden [5]. European studies using the MCOD approach revealed similar patterns in France and Italy [3], and a comprehensive Australian analysis found that some grouped conditions were mentioned on death certificates up to 35 times more often than they were selected as the underlying cause [2] (our finer three-character-level analysis identifies individual conditions with ratios exceeding 35, as the broader groupings used by Bishop et al. average across heterogeneous sub-conditions). A systematic review of MCOD methods identified rapidly growing interest in this field, with 76% of published MCOD studies published from 2001 onward, and most employing an “any-mention” approach that counts all certificate appearances of a condition [6]. Despite this growth, most studies have focused on selected conditions or disease groups rather than providing a comprehensive comparison across the full spectrum of ICD-10 codes.

The conditions most affected by undercounting have been documented individually. Hypertension is reported on Australian death certificates in only a fraction of decedents with documented hypertensive disease [7], and diabetes appears as the underlying cause in fewer than 20% of deaths where it is mentioned anywhere on the certificate [8,9]. Chronic kidney disease shows a similarly large discrepancy between underlying cause counts and total certificate mentions, a pattern observed internationally [10]. For psychiatric disorders, MCOD analysis has revealed substantial cross-country differences in how mental health conditions are recorded and coded, with most deaths involving mental illness attributed to other underlying causes [11]. Death records also provide valuable information about multimorbidity patterns at the population level, information that is lost when only the underlying cause is analysed [12]. These individual-condition studies collectively demonstrate that undercounting is widespread, but no study has quantified undercounting at the full three-character ICD-10 level to produce a comparable ranking.

This gap has practical consequences. The Global Burden of Disease Study [13] and national burden-of-disease estimates [14] rely primarily on underlying cause of death data for their mortality components, potentially underestimating the population impact of conditions with a high hidden burden. Recent Australian work has begun incorporating multiple causes into years of life lost calculations [15], but the degree of undercounting has not been quantified at the full granularity of the ICD-10 classification or systematically linked to healthcare utilisation data.

The ratio of multiple cause mentions to underlying cause deaths is an established indicator in the MCOD literature, formalized as the Standardized Ratio of Multiple to Underlying cause (SRMU) by Desesquelles et al. [3]. In Australia, Bishop et al. computed the SRMU for 136 grouped cause-of-death categories using age-standardized rates, identifying hypertension (SRMU = 35.5) and renal failure (SRMU = 8.7) as the most undercounted conditions [2]. However, these analyses used broad cause groupings rather than individual ICD-10 codes, and did not link the ratio to healthcare utilisation or prescribing data.

We adopt this established concept in its crude (non-age-standardised) form. The ABS does not currently publish age-stratified Cube 10 data, making proper age-standardisation infeasible at the three-character ICD-10 level. We therefore compute a **crude SRMU**: the raw count of death certificate mentions divided by the raw count of underlying cause deaths, computed within a single population-year. We refer to this simply as “the ratio” or “multiple-to-underlying ratio” (abbreviated MUR in figures and tables) throughout. The limitation is that the crude ratio may be inflated for conditions concentrated in older decedents, who accumulate more comorbidities; we address this through sensitivity analyses.

A ratio of 1.0 indicates a condition is mentioned only when it is the underlying cause: nothing is hidden. A ratio of 10 means the condition appears on death certificates 10 times more often than it is counted in underlying cause statistics, with 90% of its death certificate burden invisible to standard reporting (the hidden fraction is 1 − 1/ratio). To illustrate: if a condition is recorded as the underlying cause on 100 death certificates but mentioned anywhere on 2,800 certificates, its ratio is 28, its hidden fraction is 1 − 1/28 = 96%, and standard statistics would report 100 deaths while 2,700 additional deaths where it contributed go uncounted. A condition like suicide by hanging (ratio ≈ 1.0), on the other hand, appears on death certificates almost exclusively as the underlying cause, because the mechanism of death *is* the initiating cause, so there is no hidden burden (Figure 1).

**Figure 1:**
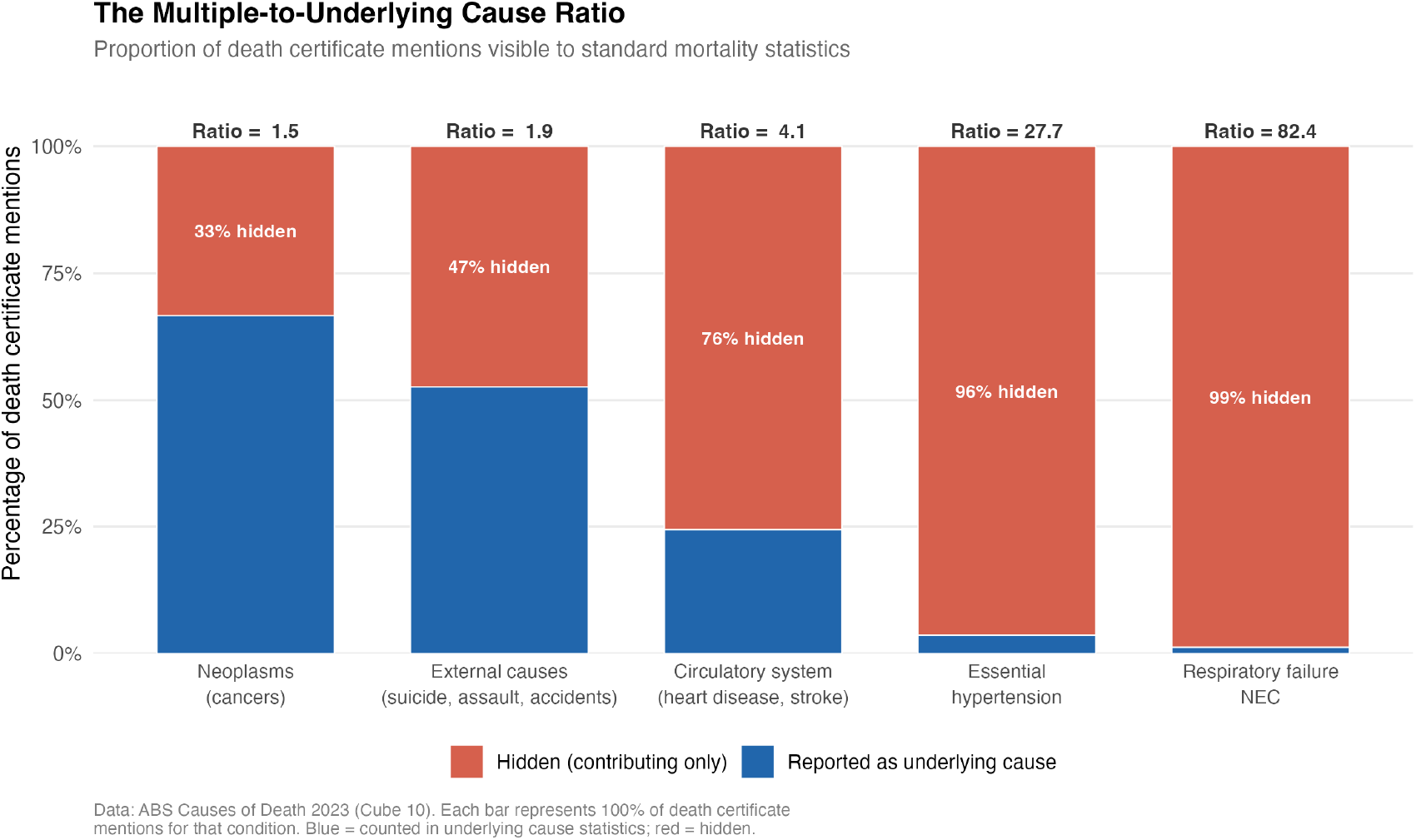
Conceptual illustration of the multiple-to-underlying ratio (MUR). Stacked bars show the proportion of death certificate mentions reported as the underlying cause (blue, visible to standard statistics) versus contributing only (red, hidden). Five conditions spanning the full ratio range are shown. Data: ABS Causes of Death 2023, Cube 10.

**Figure 2:**
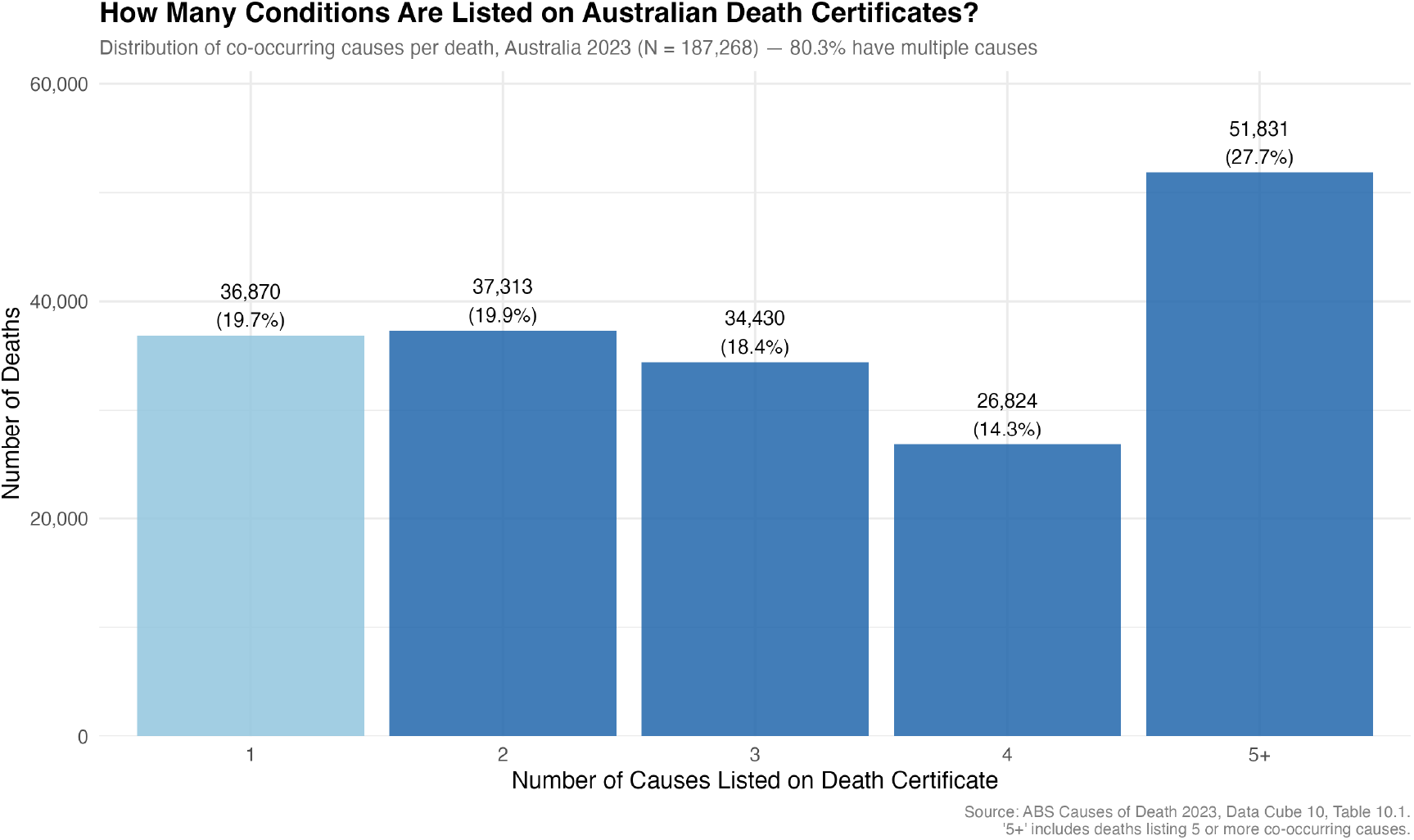
Distribution of the number of causes per death certificate (N = 187,268). The average of 3.5 causes means the underlying cause represents only ∼29% of recorded morbidity at death.

**Figure 3:**
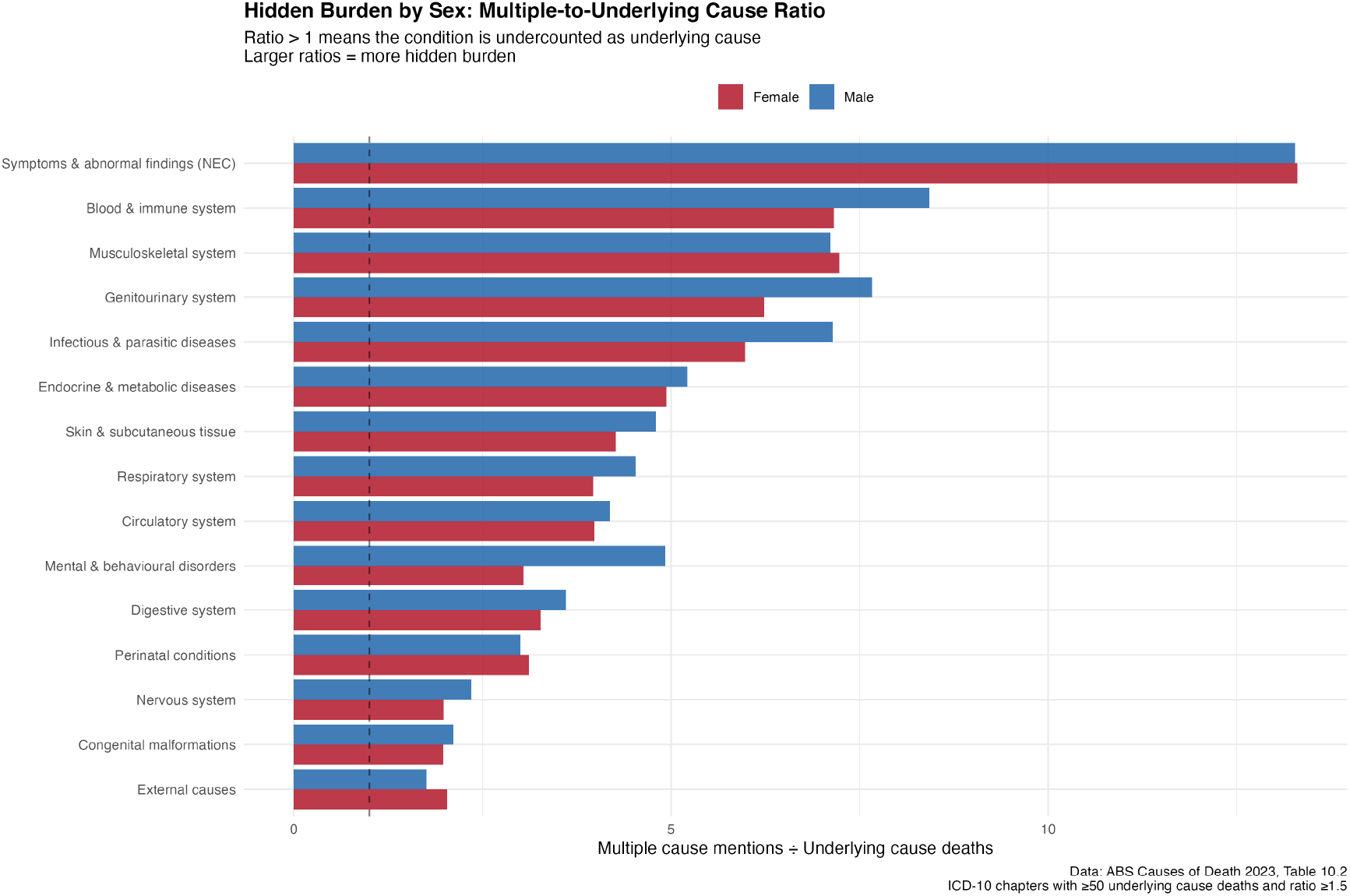
Sex-stratified multiple-to-underlying ratio (MUR) by ICD-10 chapter. Males consistently show higher ratios across most disease chapters.

**Figure 4:**
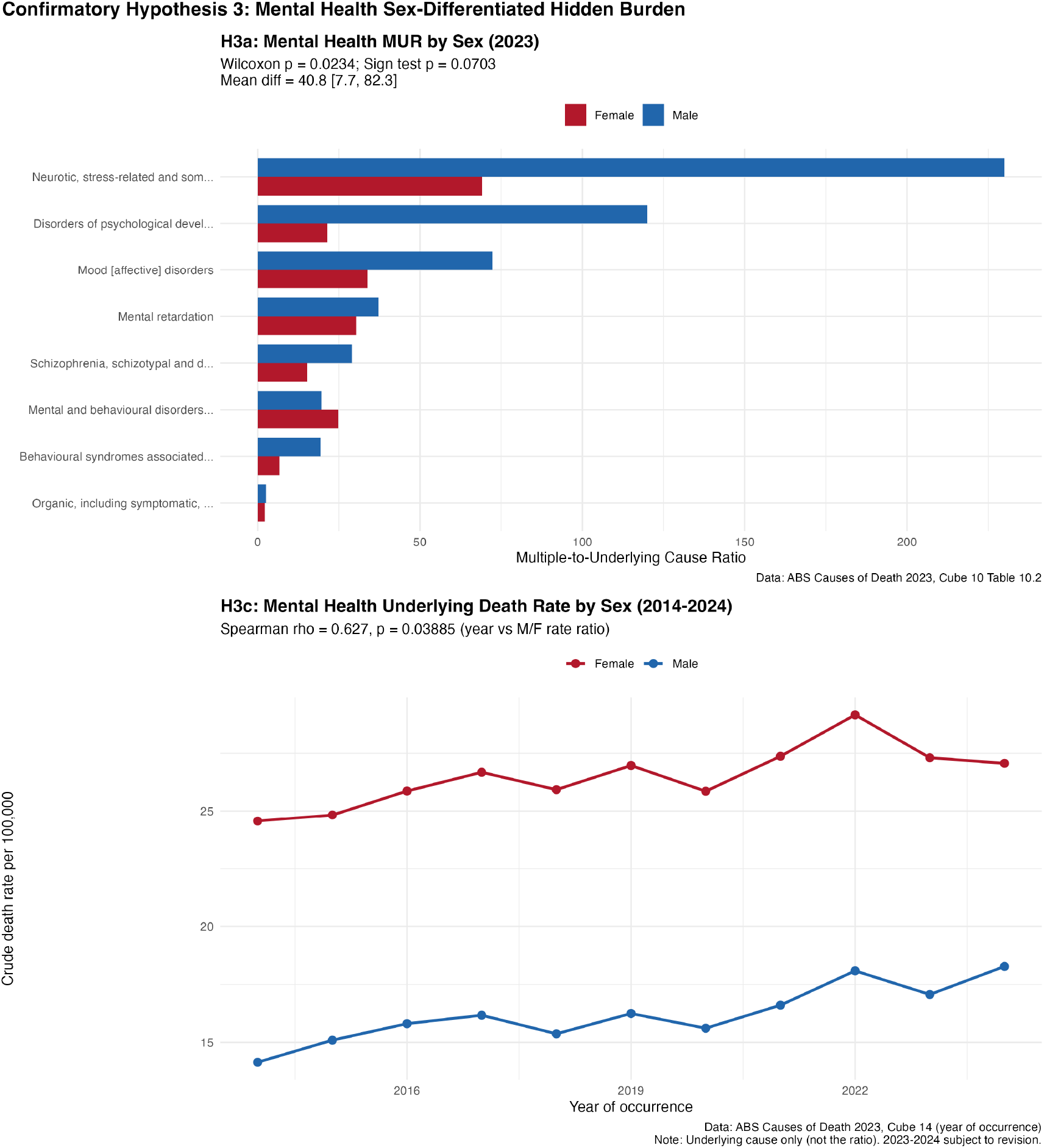
Mental health ratio by sex (Hypothesis H3). Seven of eight block-level conditions showed higher male than female ratios (uncorrected p = 0.023, effect size r = 0.80).

**Figure 5:**
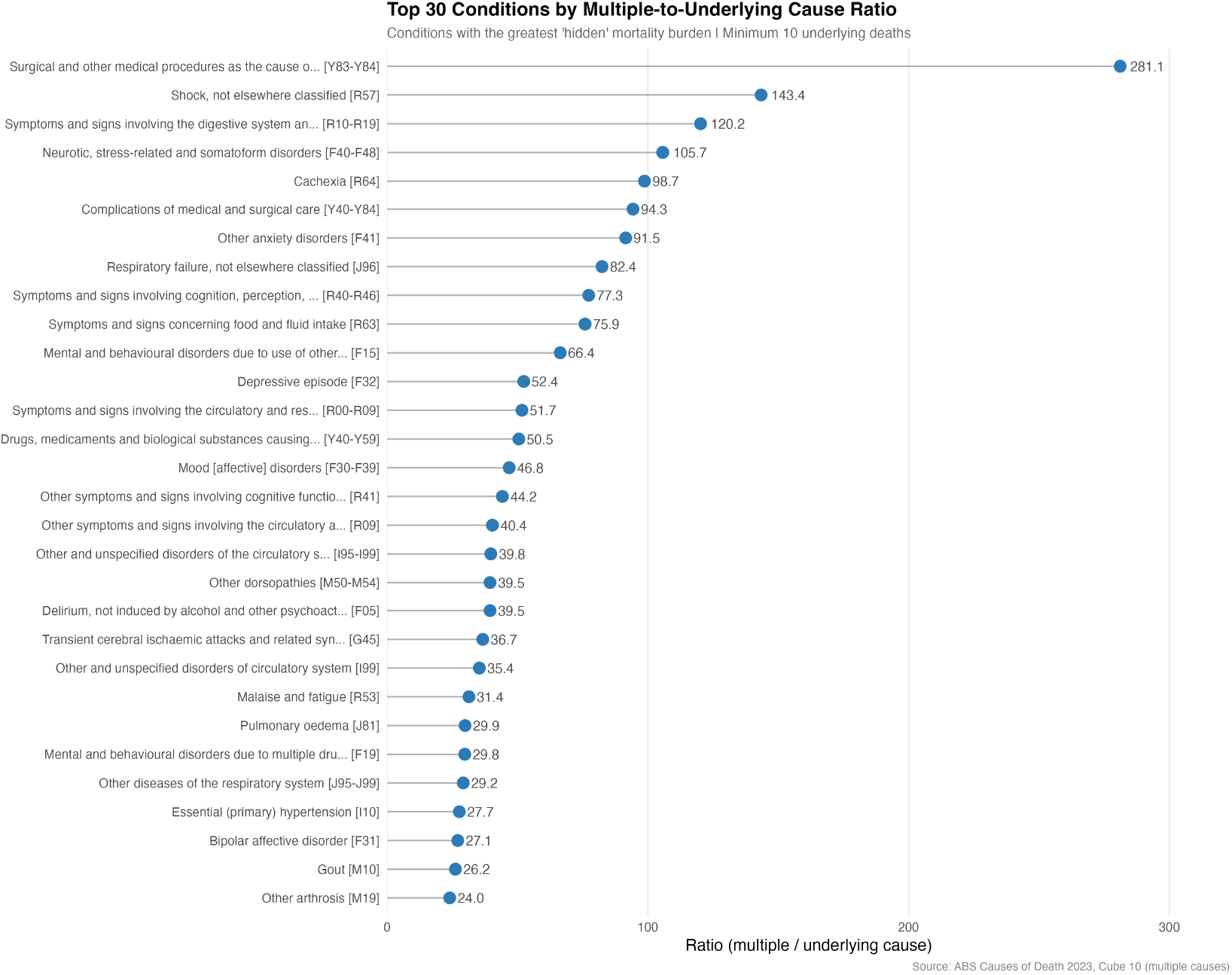
Ratio ranking for the top 30 conditions (Cleveland dot plot). The highest-ratio conditions cluster in symptom/sign codes, mental health, and complication codes.

We use “hidden” in a technical sense throughout. These conditions are recorded by certifying physicians and present in the raw data, but absent from the summary statistics that inform most mortality reports. The underlying cause convention serves a legitimate function; the ratio simply quantifies what is lost by that necessary simplification.

We aimed to compute and rank the multiple-to-underlying ratio for all ICD-10 conditions at the three-character code level in Australian mortality data, extending previous grouped analyses [2] to the full granularity of the classification. We also tested three pre-registered hypotheses concerning sex differences and geographic variation in hidden burden, and examined sex-stratified patterns across the disease spectrum.

## Methods

### Study design and pre-registration

This was a cross-sectional analysis of publicly available, aggregated administrative data. The study was pre-registered on the Open Science Framework (OSF) prior to confirmatory analysis (https://doi.org/10.17605/OSF.IO/K46RN). Three confirmatory hypotheses were specified with exact statistical tests and a Holm-Bonferroni correction plan. Exploratory analyses were also pre-specified as a separate category. Reporting follows the RECORD (REporting of studies Conducted using Observational Routinely-collected health Data) extension to STROBE [26], appropriate for studies using routinely-collected administrative data. No ethics approval was required as all data are publicly released in aggregated, de-identified form.

### Data sources

### Multiple causes of death

The Australian Bureau of Statistics (ABS) Causes of Death 2023 data (released 2024) [16] includes two relevant cubes. Cube 10 provides counts of deaths where each ICD-10 condition was recorded as (a) the underlying cause and (b) any cause (multiple cause count), stratified by sex; this formed the basis for all ratio calculations. Cube 10 is publicly released as aggregated counts by condition and sex, without age stratification at the condition level. Unit record files that would enable direct age-standardisation at the three-character ICD-10 level are restricted-access microdata not available for this analysis; this is the principal reason the ratio is computed in crude form. Cube 14 provides underlying cause deaths by year of occurrence (2014–2024, subject to revision for recent years) and was used for exploratory temporal trend analyses. Data were available for a single cross-sectional year for multiple cause analysis (deaths registered in 2023, which includes most deaths occurring in 2023 plus late registrations from 2022).

### Avoidability classification

Conditions were classified as potentially avoidable (preventable or treatable) versus non-avoidable using the AIHW National Healthcare Agreement Performance Indicator 16 definitions [17].

### Population estimates

Mid-year (June quarter) estimated resident populations by state/territory and sex from the ABS [18] were used for rate calculations.

### External validation data

US Centers for Disease Control and Prevention (CDC) WONDER Multiple Cause of Death (D77) and Underlying Cause of Death (D76) databases for 2020 [19] were used to validate age-standardisation effects. These databases provide age × cause cross-tabulations that are not available from Australian sources.

### Ratio computation

For each ICD-10 condition *i*, the multiple-to-underlying ratio was calculated as:

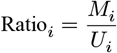

where M_i is the multiple cause count (number of deaths where condition *i* appeared anywhere on the death certificate) and U_i is the underlying cause count (number of deaths where condition *i* was selected as the underlying cause). The percentage of “hidden” deaths was calculated as (1 − 1/ratio) × 100. Sex-specific ratios were computed analogously using sex-specific numerators and denominators. The ratio was calculated at the three-character ICD-10 code level, the block level (grouped codes, e.g., I10–I15), and the chapter level (e.g., all diseases of the circulatory system, I00–I99).

Two thresholds were applied: conditions with fewer than 10 underlying cause deaths (persons) were excluded from all ranking analyses, and conditions with fewer than 50 underlying deaths were flagged as having unstable estimates with wide confidence intervals. The ≥10 threshold ensures a minimum denominator for ratio computation; the ≥50 threshold distinguishes between conditions where the ratio point estimate is reliable and those where it carries substantial statistical uncertainty.

### Important interpretive caveat

Unlike the age-standardised SRMU used by Bishop et al. [2], the crude ratio uses raw counts without age adjustment. This means conditions that predominantly kill older people may have mechanically inflated ratios, because older decedents have more conditions recorded on their death certificates (averaging ∼4.5 causes at ages 85+ versus ∼2.0 at ages 20–40) [2,12]. The ratio ranking should therefore be interpreted cautiously for cross-condition comparisons: a high ratio may partly reflect that a condition’s decedents are older (and thus have more multi-morbidity) rather than that the condition is more “hidden.” Comparisons within age-similar disease groups are more valid than comparisons across conditions with very different age-at-death profiles. A sensitivity analysis (described below) was performed to assess the magnitude of this potential confound.

A related caveat is that not all certificate mentions necessarily reflect causal contribution to death. A condition listed in Part II (“other significant conditions”) may range from a major contributor (e.g., diabetes impairing immune function in a sepsis death) to a clinically incidental comorbidity (e.g., well-controlled hypertension listed for an elderly cancer death where it played no causal role). The ratio treats all mentions equally and therefore captures *administrative prevalence on death certificates* rather than confirmed causal contribution. This “bystander” effect likely inflates the ratio for highly prevalent chronic conditions such as hypertension, which certifiers may list routinely as a comorbidity regardless of its role in the specific death. The ratio should thus be interpreted as an upper bound on the true hidden burden, with the degree of overestimation varying by condition.

### Confirmatory hypotheses

Three hypotheses were pre-registered (OSF numbering):

#### H1 (Hypertension sex-differentiated hidden burden)

We hypothesised that the multiple-to-underlying ratio for hypertensive diseases (I10–I15) would differ between males and females. The pre-registered test was a paired Wilcoxon signed-rank test across 17 years of ratio data. Because ABS Cube 10 was available for only a single year (2023), we adapted the test to use the four hypertension sub-conditions (I10–I13) as paired observations, applying the same non-parametric test.

#### H2 (Geographic variation by preventability)

We hypothesised that potentially avoidable conditions would show greater geographic variation in mortality rates (measured by the coefficient of variation [CV] across eight states and territories) than non-avoidable conditions. The pre-registered test was a Mann-Whitney U test comparing CVs.

#### H3 (Mental health sex-differentiated hidden burden)

We hypothesised that the multiple-to-underlying ratio for mental and behavioural disorders (F00–F99) would differ between males and females, driven by substance use disorders (F10–F19). The pre-registered test mirrored H1, adapted to use eight block-level sub-conditions as paired observations.

#### Multiple testing correction

We applied Holm-Bonferroni correction across the three primary tests (one per hypothesis family). Secondary tests within each family were conditional on the primary test achieving significance.

### Deviations from pre-registration

The principal deviation was the unit of analysis for H1 and H3. The pre-registration specified year-level ratio pairs (n = 17 years), but the ABS 2023 data (released 2024) contained Cube 10 for a single year only, precluding multi-year ratio computation. We adapted to use sub-conditions within the disease group as paired observations (n = 4 for H1, n = 8 for H3), preserving the non-parametric test structure but reducing statistical power. Beyond reducing power, this adaptation changed the unit of inference from temporal replication (the same ratio measured across years) to within-group replication (the sex difference measured across related but distinct conditions). The adapted tests therefore evaluate whether the sex difference generalises across sub-conditions within a disease group, rather than whether it is temporally stable, a related but conceptually distinct question. Replication with multi-year data remains necessary to answer the original pre-registered question. This deviation is transparently reported.

For H2, age-standardised rates (ASR) were used where the ABS published them; crude rates were used for conditions where ASR was suppressed due to small numbers in individual jurisdictions. The rate type is recorded per condition.

#### Note on the role of confirmatory tests

The primary contribution of this study is descriptive: a systematic quantification of hidden mortality burden across all ICD-10 conditions. The three pre-registered hypotheses were intended as illustrative tests of specific predictions about the ratio distribution, not as the study’s central claims. Because data limitations necessitated adapting the tests (reducing n from 17 years to 4–8 sub-conditions), the formal hypothesis tests should be interpreted as pre-specified analyses that could not be executed as designed, rather than as definitive confirmatory evidence. The descriptive patterns (effect sizes, confidence intervals, and consistency across sub-conditions) provide the primary evidence for evaluating the hypothesized relationships.

### Exploratory analyses

Seven exploratory analyses were pre-specified (E1–E7). Two additional exploratory analyses were conducted post hoc:

- **E-HM4:** Full ratio ranking supplementary table with sex stratification.
- **E-HM5:** Counterfactual analysis quantifying the contribution of suicide coding rules to the male mental health ratio excess.

All exploratory results are reported without multiple testing correction and are clearly labelled as hypothesis-generating.

### Age-adjustment sensitivity analysis

Because the ABS does not currently publish age-stratified multiple cause data at the three-character ICD-10 level, direct age standardisation was not feasible. We therefore performed a sensitivity analysis using two complementary approaches:

#### Method 1 (Indirect adjustment)

For each condition, we estimated the expected ratio based on the median age at death and the known relationship between age and certificate complexity (approximately 2.0 causes per certificate at ages 0–20 increasing to 4.5 causes at ages 85+) [2,12]. The ratio of observed to expected indicates whether a condition’s hidden burden exceeds what would be predicted from its age profile alone.

#### Method 2 (Regression-based adjustment)

We modelled log(ratio) as a function of estimated median age at death across all conditions with ≥50 underlying cause deaths. The R^2^ from this regression quantifies how much of the variation is explained by age alone, and the residuals identify conditions with high ratios after accounting for age effects.

Both methods use chapter-level median age estimates as proxies for condition-specific age-at-death profiles, with refinements for conditions known to have distinctive age distributions (e.g., suicide, perinatal conditions, dementia).

#### Method 3 (External validation using US data)

As an external validation, we computed both crude and directly age-standardised ratios using US CDC WONDER Multiple Cause of Death data (2020), which provides age × cause cross-tabulations not available from Australian sources. Direct age standardisation used the US 2000 Standard Population (Census P25-1130) across 11 age groups. We compared crude versus age-standardised ratios for eight key cause groups (diabetes, dementia/Alzheimer, hypertensive diseases, ischaemic heart disease, cerebrovascular disease, chronic lower respiratory disease, renal failure, influenza/pneumonia) to assess whether the divergence exceeds a 10% materiality threshold.

These sensitivity analyses cannot fully replicate age-standardised SRMU computation [2,3] but provide a practical assessment of whether the ratio ranking is robust to age confounding.

### Statistical analysis

All analyses were performed in R (version 4.5.2) using the tidyverse, coin, readxl, and scales packages. Confidence intervals for ratios (Table 1) were computed using exact Poisson confidence intervals for the underlying cause count, propagated to the ratio scale. Because these are complete-population counts for a single registration year rather than a sample, the Poisson model captures year-to-year registration variability rather than sampling uncertainty; the multiple cause count (numerator) is treated as fixed given the denominator. Spearman rank correlations were used for monotonic associations. Pearson correlations were used for log-transformed variables where linearity was established. Trend classification for temporal data used Spearman rho (threshold |rho| > 0.3 and p < 0.10). Bootstrap confidence intervals used 10,000 resamples with bias-corrected and accelerated (BCa) intervals where bootstrap skewness supported their use (H1), and standard percentile intervals otherwise (H3); however, with very small sample sizes (n = 4 for H1, n = 8 for H3), bootstrap CIs are at the edge of reliability and should be interpreted cautiously as indicative rather than definitive.

**Table 1.** Highest-ratio conditions by multiple-to-underlying ratio, Australia 2023.

Panel A: Stable estimates ( $\geq 50$ underlying deaths)
| Rank | Condition | ICD-10 | Level | Underlying | Multiple | Ratio | 95% CI | % Hidden |
| --- | --- | --- | --- | --- | --- | --- | --- | --- |
| 1 | Complications of medical and surgical care | Y40–Y84 | Block | 68 | 6,413 | 94.3 | 74.3–121.8 | 98.9% |
| 2 | Respiratory failure, not elsewhere classified | J96 | 3-char | 92 | 7,582 | 82.4 | 67.2–102.2 | 98.8% |
| 3 | Cognitive, perceptual and emotional symptoms | R40–R46 | Block | 55 | 4,250 | 77.3 | 59.4–101.9 | 98.7% |
| 4 | Depressive episode | F32 | 3-char | 71 | 3,723 | 52.4 | 41.5–67.1 | 98.1% |
| 5 | Circulatory and respiratory symptoms | R00–R09 | Block | 75 | 3,876 | 51.7 | 41.2–65.7 | 98.1% |

Panel B: Higher point estimates but unstable ( $< 50$ underlying deaths)
| Rank | Condition | ICD-10 | Level | Underlying | Multiple | Ratio | 95% CI | % Hidden |
| --- | --- | --- | --- | --- | --- | --- | --- | --- |
| - | Surgical and medical procedure complications | Y83–Y84 | Block | 15 | 4,217 | 281.1 | 170.5–502.3 | 99.6% |
| - | Shock, not elsewhere classified | R57 | 3-char | 24 | 3,442 | 143.4 | 96.7–222.3 | 99.3% |
| - | Digestive symptoms and signs | R10–R19 | Block | 20 | 2,403 | 120.2 | 78.0–197.6 | 99.2% |
| - | Neurotic and stress-related disorders | F40–F48 | Block | 22 | 2,325 | 105.7 | 70.0–170.6 | 99.1% |
| - | Other anxiety disorders | F41 | 3-char | 19 | 1,739 | 91.5 | 58.6–153.0 | 98.9% |

*Note: Panel A shows conditions with stable estimates ( $\geq 50$ underlying deaths) and relatively narrow confidence intervals; these are the robust findings. Panel B shows conditions with higher point estimates but small denominators; their wide CIs (e.g., 170–502 for Y83–Y84) indicate substantial uncertainty in the precise magnitude. The “Level” column indicates whether each entry is a single three-character ICD-10 code (3-char) or a block-level grouping of related codes (Block). Y83–Y84 (Panel B) is a subset of Y40–Y84 (Panel A); the subset has a higher ratio because it isolates surgical and medical procedure complications (denominator $n = 15$ ) from the broader block that includes drug and biological adverse effects (denominator $n = 68$ ).*
Conditions with ratio = 1.0 were exclusively external causes (suicide methods, assault, undetermined intent events) where the mechanism of death is inherently the underlying cause.

## Results

### Descriptive: Death certificate complexity

In 2023, 187,268 deaths were registered in Australia (year of registration, not year of occurrence) with complete cause-of-death information available for ratio analysis. An average of 3.5 conditions were recorded per death certificate, consistent with international estimates of 3–4 causes per certificate [2,4]. Only 19.7% of deaths listed a single cause; 80.3% reported two or more causes. The distribution peaked at 3–4 causes per death, with a long right tail extending to 15+ causes (Supplementary Figures S3 and S4).

### Ratio distribution

Among 663 conditions with at least 10 underlying cause deaths, the multiple-to-underlying ratio ranged from 1.0 to 281.1 (Table 1). The distribution was highly right-skewed: median ratio was 2.5, mean was 6.6, and 71 conditions (10.7%) had a ratio exceeding 10. Among conditions with stable estimates (≥50 underlying deaths; Table 1, Panel A), complications of medical care (Y40–Y84, ratio = 94.3) had the highest value, followed by respiratory failure (J96, ratio = 82.4) and cognitive symptoms (R40–R46, ratio = 77.3). Respiratory failure’s extreme ratio reflects WHO coding rules that classify it as a “mode of dying” rather than an initiating cause; certifiers are instructed not to select terminal events as the underlying cause [1], so it appears on over 7,500 certificates but is selected as underlying cause for only 92. Several conditions with fewer than 50 underlying deaths showed even higher point estimates (Table 1, Panel B), but these carry substantial statistical uncertainty: the CI for surgical complications (Y83–Y84) spans 170 to 502.

Conditions with ratio = 1.0 were exclusively external causes (suicide methods, assault, undetermined intent events) where the mechanism of death is inherently the underlying cause.

### Chapter-level ratios

At the ICD-10 chapter level (Table 2), the highest ratio was observed for diseases of the eye and adnexa (ratio = 76.5; 13 underlying deaths, 995 multiple cause mentions), followed by diseases of the ear (ratio = 15.1) and symptoms/signs not elsewhere classified (ratio = 13.3). Neoplasms had the lowest chapter-level ratio (1.5), reflecting the ICD coding convention that malignant neoplasms, once identified as initiating the fatal sequence, take priority as the underlying cause [1]. Supplementary Figure S1 shows the absolute (rather than proportional) hidden burden by chapter.

**Table 2.** Chapter-level multiple-to-underlying ratios, ranked by descending ratio.

| ICD-10 Chapter | Underlying deaths | Multiple cause mentions | Ratio | % Hidden |
| --- | --- | --- | --- | --- |
| Eye and adnexa | 13 | 995 | 76.5 | 98.7% |
| Ear and mastoid process | 19 | 287 | 15.1 | 93.4% |
| Symptoms, signs, abnormal findings | 3,781 | 50,243 | 13.3 | 92.5% |
| Blood and immune disorders | 634 | 4,924 | 7.8 | 87.1% |
| Musculoskeletal system | 1,820 | 13,080 | 7.2 | 86.1% |
| Genitourinary system | 5,002 | 34,551 | 6.9 | 85.5% |
| Infectious and parasitic diseases | 3,108 | 20,367 | 6.6 | 84.7% |
| Endocrine, nutritional, metabolic | 8,514 | 43,299 | 5.1 | 80.3% |
| Skin and subcutaneous tissue | 780 | 3,516 | 4.5 | 77.8% |
| Respiratory system | 17,088 | 72,765 | 4.3 | 76.5% |
| Circulatory system | 42,298 | 173,121 | 4.1 | 75.6% |
| Mental and behavioural disorders | 12,337 | 46,893 | 3.8 | 73.7% |
| Digestive system | 7,958 | 27,440 | 3.4 | 71.0% |
| Nervous system | 12,544 | 27,115 | 2.2 | 53.7% |
| Congenital malformations | 650 | 1,333 | 2.1 | 51.2% |
| External causes | 13,307 | 24,664 | 1.9 | 46.0% |
| Neoplasms | 52,819 | 76,987 | 1.5 | 31.4% |

*Note: Three ICD-10 chapters are omitted: pregnancy/childbirth (5 underlying deaths), injury/poisoning (0 underlying deaths, as nature-of-injury codes are not selected as underlying cause under ICD-10 coding rules [1]), and perinatal conditions (566 underlying deaths, ratio = 3.1; omitted because perinatal mortality has a distinct epidemiological profile that makes ratio comparisons with other chapters less meaningful). Confidence intervals are not shown for chapter-level ratios because chapters aggregate many heterogeneous conditions, making a single Poisson CI misleading. Underlying death counts range from 13 (eye and adnexa) to 52,819 (neoplasms); ratios for chapters with small denominators, particularly eye and adnexa (n = 13) and ear (n = 19), carry substantial uncertainty and should be interpreted cautiously. Condition-level CIs are provided in Table 1.*

### Sex-stratified ratios

Males consistently showed higher ratios than females across most conditions, indicating a greater relative hidden burden in male mortality (Supplementary Figure S6). Among the 15 conditions with ratio > 2 and at least 10 underlying deaths in both sexes where the male-to-female ratio exceeded 2.0, the most extreme sex divergence was observed for intestinal malabsorption (K90; male/female ratio = 5.73), other anxiety disorders (F41; ratio = 4.63), and gastritis and duodenitis (K29; ratio = 4.50). The full list of sex-divergent conditions is provided in Supplementary Table S2.

### Age-adjustment sensitivity analysis results

The regression-based adjustment (Method 2) yielded R^2^ = 0.109, indicating that **only 10.9% of the variation in the ratio across conditions is explained by estimated median age at death**. When raw ratio rankings were compared to age-adjusted rankings using both methods, no conditions were identified as primarily “age-driven” (i.e., in the top 30 on raw ratio but dropping below rank 50 after adjustment). The top 15 high-ratio conditions retained their approximate rankings after both adjustment methods (Supplementary Table S6; Supplementary Figures S13–S14). The R^2^ across all conditions is complemented by this direct rank-stability check for the top conditions specifically, since the concern about age confounding applies most acutely to the highest-ranked entries. Together, these findings support the validity of the ratio *ranking* for identifying conditions with elevated hidden burden: the relative ordering is robust, even though absolute values may be approximate.

The external validation (Method 3) using US CDC WONDER data showed that **6 of 8 cause groups had material divergence** (>10%) between crude and age-standardised ratios. Only chronic lower respiratory diseases (+0.6%) and ischaemic heart disease (+7.1%) showed concordant estimates. For the remaining causes, age standardisation substantially reduced the ratio for conditions concentrated in the elderly: diabetes (crude 3.80 vs age-standardised 2.49, −34%), hypertensive diseases (crude 12.84 vs age-standardised 10.33, −20%), and influenza/pneumonia (crude 5.98 vs age-standardised 4.98, −17%). Conversely, cerebrovascular disease (+17%), renal failure (+16%), and Alzheimer disease (+21%) showed higher age-standardised than crude ratios. Despite these absolute-value shifts, the rank ordering was nearly perfectly preserved: the Spearman rank correlation between crude and age-standardised rankings across the eight cause groups was ρ = 0.976, with only one adjacent rank swap (influenza/pneumonia and renal failure exchanging positions 2 and 3). This indicates that the crude ratio preserves the ranking of high-ratio conditions but absolute values should be interpreted as approximate, with divergence of 16–34% in either direction depending on the condition. Notably, the direction did not follow a simple age-concentration pattern: Alzheimer disease, among the most elderly-concentrated conditions analysed, showed a *higher* age-standardised ratio (+21%), likely because age standardisation shifts denominators toward younger age groups where underlying cause deaths are disproportionately rare relative to mentions. The direction of divergence thus depends on the relative age distribution of underlying versus multiple cause counts, not simply on median age at death. A proper age-standardised SRMU at three-character level would require age-stratified Cube 10 data that the ABS does not currently publish.

### Confirmatory hypothesis results

#### H1: Hypertension sex-differentiated hidden burden

The aggregate ratio for hypertensive diseases (I10–I15) was 11.1 for males and 8.3 for females (sex ratio 1.34). All four sub-conditions showed higher male than female ratios, with essential hypertension (I10) showing the largest absolute difference (male ratio = 41.8, female ratio = 21.2; Supplementary Figure S5). The descriptive evidence consistently supports a sex difference: the effect size was large (r = 0.77), the bootstrap 95% confidence interval for the mean ratio difference (male − female) was 0.24 to 15.71 (excluding zero), and the direction was unanimous across all four sub-conditions. However, the data were insufficient for formal inferential testing. With only n = 4 paired observations, the minimum achievable two-tailed p-value for a Wilcoxon signed-rank test is 0.125, meaning the test was structurally incapable of reaching α = 0.05 regardless of effect magnitude or consistency. The observed p = 0.125 (Holm-Bonferroni corrected p = 0.25) therefore represents the strongest possible result for this sample size, not an ambiguous null (Supplementary Figure S10; Supplementary Table S4a).

#### H2: Geographic variation by preventability

Among 251 conditions classifiable as avoidable (n = 100) or non-avoidable (n = 151), the median CV was nearly identical: 30.8% for avoidable and 31.1% for non-avoidable conditions (Mann-Whitney U = 7,459, p = 0.872, rank-biserial r = 0.012). A sub-analysis comparing preventable (median CV = 30.1%) and treatable (median CV = 32.6%) conditions was also non-significant (p = 0.832). Results were robust to excluding the Northern Territory and ACT (jurisdictions with small populations and high CV) (Supplementary Figure S11; Supplementary Tables S3 and S4b). These data provide no evidence for the hypothesized geographic variation.

#### H3: Mental health sex-differentiated hidden burden

The aggregate ratio for mental and behavioural disorders (F00–F99) was 4.9 for males and 3.0 for females (sex ratio 1.62). Seven of eight block-level conditions showed higher male than female ratios, with a large effect size (r = 0.80) and a bootstrap 95% CI for the mean difference that excluded zero (7.69 to 82.33). The paired Wilcoxon test was significant at the uncorrected level (Z = 2.24, p = 0.023) but did not survive Holm-Bonferroni correction (corrected p = 0.070). With n = 8 pairs, the minimum achievable two-tailed p-value is 0.008, so significance was theoretically possible but the test remained underpowered for detecting the pre-specified effect. Contrary to the pre-registered prediction, the sex difference was driven by *non-substance* mental disorders (F20–F99; p = 0.031, r = 0.88) rather than substance use disorders (F10–F19; p = 0.625). The effect sizes and confidence intervals suggest a real sex difference that the adapted test lacked power to confirm. As with H1, the adapted test evaluates within-group generalisation (across block-level sub-conditions) rather than temporal stability (the original pre-registered question); the practical impact is smaller here because within-group consistency was strong (7 of 8 blocks in the predicted direction). Full test results and sensitivity analyses are reported in Supplementary Tables S3 and S4c.

### Exploratory analysis E-HM4: Full ratio ranking

The complete ratio ranking for 663 conditions is provided in Supplementary Table S1, with sex-stratified rankings shown in Supplementary Figure S12. Key distributional findings: 71 conditions (10.7%) had ratios > 10 (more than 90% of their death certificate burden hidden); 267 conditions (40.3%) had ratios between 2.0 and 10; and 325 conditions (49.0%) had ratios < 2.0. The highest-ratio conditions clustered in three categories: symptom/sign codes (R chapter), mental and behavioural disorders (F chapter), and complication/external cause codes (Y chapter).

### Exploratory analysis E-HM5: Suicide coding and the male mental health ratio excess

The 62% higher male ratio for mental health disorders could partly reflect the interaction between psychiatric illness and suicide mortality. In Australia, approximately three-quarters of suicide deaths are male (3.25 males per female in 2023) [16]. When a death is classified as suicide, WHO coding rules assign the underlying cause to External Causes (X60–X84) [1], while contributing mental health conditions enter only the multiple cause count, thereby inflating the ratio for psychiatric conditions.

To quantify this effect, we performed a counterfactual analysis testing a range of assumed psychiatric mention rates on suicide death certificates (30–70%), with a central estimate of 50% based on US surveillance data [20]. Removing suicide-associated psychiatric mentions reduced the male/female sex ratio from 1.62 to 1.54–1.58 depending on the assumed mention rate. This implies **the suicide coding mechanism explains a minority of the male ratio excess (sensitivity range: 6–15% across plausible assumption values)**, with the central estimate at approximately 11%. The relationship between assumed mention rate and explained fraction is approximately linear (∼2 percentage points per 10% increase in mention rate). Australian coronial death certificates may have higher psychiatric mention rates than US data suggest, because Australian deaths from external causes are investigated by coroners who routinely commission psychological autopsies and toxicology reports, potentially yielding more comprehensive psychiatric documentation than standard US medical examiner certifications. Even extrapolating to a 90% mention rate (a plausible upper bound for the Australian coronial system), the mechanism would explain approximately 19% of the sex difference. At the theoretical maximum of 100% (every suicide certificate mentioning a psychiatric condition), the explained fraction reaches approximately 21%. The remaining ∼80% must reflect other factors regardless of the assumed mention rate. Full results are in Supplementary Table S5.

## Discussion

### What the ratio reveals about Australian mortality

This study provides the most granular mapping to date of the hidden mortality burden across ICD-10 conditions in Australia, extending previous grouped SRMU analyses [2] to 663 individual three-character codes. The overarching finding is that the hidden burden is both larger and more unevenly distributed than underlying cause statistics suggest. The condition at the median ratio has 60% of its death certificate mentions hidden from underlying cause statistics (ratio = 2.5), but the distribution is highly skewed: the top decile of conditions has over 90% of mentions hidden (ratio > 10), while external causes show virtually no hidden burden (ratio ≈ 1).

None of the three pre-registered confirmatory hypotheses achieved statistical significance after Holm-Bonferroni correction. These analyses should be interpreted as **pre-registered hypotheses that could not be tested as designed due to data availability**, rather than as traditional confirmatory tests. The adaptation from year-level pairs (n = 17) to sub-condition pairs (n = 4 for H1, n = 8 for H3) reduced statistical power drastically and changed the unit of inference from temporal stability to within-group generalisation; the descriptive patterns (effect sizes, bootstrap CIs, direction consistency) provide the primary evidence, while the formal Wilcoxon p-values are largely uninformative given these constraints.

For H1, the minimum achievable p-value for a two-tailed Wilcoxon test with n = 4 is 0.125, meaning the test was structurally incapable of reaching significance at α = 0.05 regardless of effect magnitude. The observed p = 0.125 represents a perfect monotonic effect, not an ambiguous null. For H3 (n = 8 pairs, minimum p = 0.008), significance was theoretically possible but the test remained underpowered. In both cases, effect sizes were large (r = 0.77 and 0.80), bootstrap confidence intervals excluded zero, and the direction was consistent across all sub-conditions. These findings warrant replication when multi-year Cube 10 data become available, not only for statistical power, but to answer the original pre-registered question about temporal stability. H2 (geographic variation) showed a clear null effect with adequate power.

### Why some conditions are hidden more than others

The ratio ranking is not random; it reflects the logic of underlying cause selection as codified in ICD-10 [1]. Two distinct mechanisms drive high ratios, and it is important to distinguish them.

#### Coding-driven high ratios

Some conditions have high ratios primarily because coding rules explicitly prohibit or discourage their selection as underlying cause. Respiratory failure (J96, ratio = 82.4) is almost never coded as the underlying cause despite appearing on over 7,500 death certificates, because WHO rules treat it as a “mode of dying” rather than a cause; certifiers are instructed not to select terminal events [1]. Similarly, R-chapter symptom codes (cognitive symptoms, circulatory symptoms) and Y-chapter complication codes are deprioritised by design. For these conditions, the high ratio reflects a coding convention, not necessarily clinical complexity.

#### Clinically-driven high ratios

Other conditions have high ratios because they participate in death through chronic, multifactorial pathways rather than acute, single-cause mechanisms. Hypertension, diabetes, and mental health conditions frequently contribute to deaths where another condition (cancer, infection, external cause) is identified as initiating the fatal sequence. For these conditions, the high ratio reflects real clinical complexity and multi-morbidity burden that underlying-cause statistics obscure.

An important caveat for clinically-driven ratios is the **“bystander” effect**: not every certificate mention represents a genuine causal contribution to the death. A condition like hypertension may be listed routinely as a comorbidity for elderly decedents regardless of whether it materially contributed to the specific death. The ratio cannot distinguish between a causally relevant mention (diabetes impairing immune function in a sepsis death) and an incidental one (stable hypertension listed for completeness on a cancer death certificate). The ratio therefore provides an upper bound on the true hidden burden for prevalent chronic conditions. This overestimation is likely most pronounced for hypertension, the most common comorbidity on Australian death certificates, and less relevant for conditions like respiratory failure or cognitive symptoms whose presence on a certificate more directly implies clinical involvement near death.

The distinction matters for policy: coding-driven high ratios point toward statistical reform (better reporting conventions), while clinically-driven high ratios point toward clinical intervention (addressing underrecognised disease burden). The two mechanisms often co-occur. External causes (suicide methods, assault, accidents) have ratio ≈ 1.0 because the mechanism *is* the initiating cause, making both coding rules and clinical reality point to the same answer. Chronic conditions embedded in multi-morbidity have ratio » 1 for both reasons: they are clinically complex *and* coding rules favour more specific initiating causes.

The clinical implications are substantial. Respiratory failure (ratio = 82.4) appears on over 7,500 death certificates annually but is recorded as the underlying cause in only 92, a 98.8% hidden burden. This means underlying-cause statistics report ∼100 “deaths from respiratory failure” while ignoring 7,400 deaths where respiratory failure was judged by a certifying physician to have contributed. For burden-of-disease estimates, this suggests respiratory failure should be weighted approximately 80-fold higher than its underlying-cause count implies. Similarly, hypertension (ratio = 27.7) contributes to death approximately 28 times more often than it initiates the fatal sequence, with direct implications for evaluating cardiovascular prevention programs. The policy question is not whether to abandon underlying cause conventions (which serve international comparability) but whether burden estimates and resource allocation should incorporate multiple cause weights where the ratio reveals systematic understatement.

### Comparison with established Australian benchmarks

Our findings are broadly consistent with, and extend, the recent Australian MCOD analyses by Bishop et al. [2] and Joshy et al. [15]. Bishop et al. reported an SRMU of 35.5 for hypertensive diseases using 2006–2017 pooled age-standardised rates, compared with our single-year crude ratio of 27.7 for essential hypertension. Part of this gap reflects age confounding: US CDC validation showed age standardisation reduced the hypertension ratio by 20% (crude 12.84 vs age-standardised 10.33), with the remainder attributable to period differences and ICD-10 grouping level. Hypertension remained the top-ranked condition in both studies. Our finding of 3.5 causes per death certificate matches the 3.4 reported by Bishop et al. [2] and the international range of 3–4 causes [4,6]. The primary extension is granularity: 663 three-character codes versus 136 grouped categories, enabling identification of specific conditions with exceptionally high hidden burden (e.g., anxiety disorders F41, respiratory failure J96) that would be averaged away in broader groupings.

### Sex differences in hidden burden

The consistent pattern of higher male ratios across most conditions, particularly striking for mental health disorders (male ratio 62% higher than female), suggests that male deaths are characterised by greater multi-morbidity complexity. This aligns with evidence that Australian males have higher rates of comorbid conditions at death [21] and with the finding that death certificate complexity, as measured by the number of conditions per certificate, varies by sex and age [12]. This pattern may partly reflect later healthcare engagement and more advanced disease at the time of diagnosis among males, consistent with systematic review evidence that masculinity norms discourage men from seeking help for depression and other conditions [22].

The unexpected finding that the sex difference in mental health ratios was driven by non-substance disorders (F20–F99) rather than substance use disorders (F10–F19) challenges assumptions about the gendered nature of hidden mental health mortality. While substance use disorders are approximately twice as prevalent among males as females in Australia [23], the hidden burden, as measured by the ratio, appears more sex-differentiated for conditions like anxiety disorders (F41; male/female sex ratio = 4.63), neurotic disorders (F40–F48; sex ratio = 3.33), and bipolar disorder (F31; sex ratio = 2.39). This finding suggests that psychiatric comorbidity at death is systematically more underrecorded for males, consistent with international evidence on sex differences in psychiatric death certification [11].

One potential mechanical driver of the higher male mental health ratio is the interaction between mental illness and suicide mortality, the “suicide coding mechanism,” whereby WHO rules assign suicide deaths to External Causes while psychiatric contributors enter only the multiple cause count, artificially inflating the ratio for mental health conditions. Our counterfactual analysis (E-HM5) estimated this mechanism explains a minority of the male ratio excess (sensitivity range: 6–15% across assumed psychiatric mention rates of 30–70%). Even under the most generous assumption (that 90% of Australian suicide death certificates mention a psychiatric condition, plausible given the systematic nature of coronial investigations), the mechanism explains approximately 19% of the sex difference. The remaining ∼80% must reflect other factors: later healthcare engagement among males, more advanced disease at diagnosis, sex differences in death certification practices, or real differences in multi-morbidity burden at death.

This finding aligns with Spark et al. [24], who demonstrated empirically that underlying-cause-only analyses under-estimated mortality associated with mental health and substance use disorders by 41% in a US veteran cohort, precisely because ICD coding rules assigned external causes as the underlying cause in suicide and overdose deaths. The suicide-mediated mechanism may also partly explain the unexpected finding that the sex difference was driven by non-substance mental disorders rather than substance use: mood disorders (F30–F39) showed a male/female sex ratio of 2.14, and meta-analyses consistently find mood disorders carry an 8–11-fold increased suicide risk [25], while substance use disorders (F10–F19) showed a slight female excess (sex ratio = 0.79). Non-substance psychiatric conditions like depression and anxiety are almost never selected as the underlying cause in the presence of any physical disease or external cause [1], making them more sensitive to this coding dynamic than substance disorders, which may be certified directly as poisoning.

### Geographic variation: an unexpectedly uniform picture

Perhaps surprisingly, we found no evidence that avoidable conditions show greater geographic variation than non-avoidable ones. The near-identical CVs (30.8% vs 31.1%) suggest that geographic variation in Australian mortality is not systematically greater for conditions amenable to prevention or treatment. This null finding has several possible interpretations.

The first is that Australian health system infrastructure is sufficiently uniform across states and territories that geographic access to prevention and treatment does not produce differential mortality patterns. This interpretation aligns with Australia’s universal healthcare system (Medicare), which provides relatively uniform access to primary care across states and territories [21].

The second, more critical interpretation is that the AIHW avoidability classification [17] may not capture the dimensions of mortality most sensitive to geographic factors. The classification was developed primarily to measure health system performance, not geographic equity, and conditions like ischaemic heart disease are classified as “avoidable” despite having substantial non-modifiable components. A classification specifically designed around geographic sensitivity (incorporating remoteness, Indigenous health disparities, or socioeconomic accessibility) might yield different results.

A third possibility involves a statistical tension visible in the data. The Northern Territory shows strikingly elevated mortality across nearly all conditions (Supplementary Figure S8), a pattern driven substantially by Indigenous health disparities [21] in a jurisdiction where Indigenous Australians comprise approximately 27% of the population compared with 3.8% nationally [18]. Yet the aggregate CV comparison pools this extreme outlier with seven more homogeneous jurisdictions. Our sensitivity analysis excluding the Northern Territory and ACT (small-population jurisdictions with elevated CV) yielded similar results, but this exclusion may mask precisely the geographic variation that avoidability ought to capture. The null finding may thus reflect the limitations of state-level geographic units rather than the absence of avoidability-sensitive variation at finer spatial scales.

A fourth, methodological consideration is that the coefficient of variation across only 8 jurisdictions is itself a noisy statistic. With so few data points per condition, the CV estimate has high sampling variance, and the Mann-Whitney U test is comparing distributions of noisy CVs between two categories. This power/precision issue may contribute to the null independent of the substantive interpretations above. State-level mortality patterns are detailed in Supplementary Figures S7–S9.

### Implications for health policy and statistics

These findings have direct implications for how mortality data are used in health policy. Current burden-of-disease estimates [13,14] substantially undercount conditions with high ratios. For conditions like hypertension (ratio = 27.7) or respiratory failure (ratio = 82.4), underlying cause counts capture less than 4% and 1.2% of the death certificate burden, respectively. Incorporating multiple cause analysis into routine burden estimation would correct this imbalance, particularly for chronic conditions managed in primary and hospital care.

The multiple-to-underlying ratio also raises questions about performance indicators. Metrics based on underlying cause mortality, such as potentially avoidable death rates [17], may systematically favour conditions with high ratios, because deaths where these conditions contribute are attributed to other causes. Several approaches could address this: (1) **ratio-weighted burden estimates**, where underlying cause counts are multiplied by condition-specific ratios to approximate total death certificate burden; (2) **routine dual reporting** of both underlying and multiple cause counts in official statistics, as some European countries already practice [3]; or (3) **threshold-based flagging** that identifies conditions where the ratio exceeds a defined threshold (e.g., >10) and recommends multiple cause analysis for policy decisions involving those conditions. The consistent male excess in ratios across most conditions further argues for sex-disaggregated multiple cause analysis as routine practice. Finally, monitoring the ratio over time could provide early signals of changing multi-morbidity patterns. The current study was limited to a single year of ratio data; ongoing multiple cause releases from the ABS would enable trend analysis.

### Strengths and limitations

#### Strengths

This study analyses the complete Australian mortality dataset (187,268 deaths) rather than a sample, and computes the multiple-to-underlying ratio across the full spectrum of ICD-10 conditions rather than for selected diseases. The analysis was pre-registered, deviations are transparently reported, and all results, including null findings, are presented. The ratio is simple to compute, interpret, and replicate from publicly available data. We acknowledge that the study’s scope, spanning descriptive, confirmatory, and exploratory analyses, necessarily trades depth for breadth; a focused study of a single disease domain could pursue more granular questions, but the comprehensive approach here was chosen deliberately to establish the landscape-level view that individual studies can build on.

#### Limitations

First, and most importantly, the ratio is not age-standardised, unlike the SRMU used by Bishop et al. [2]. This matters because older decedents accumulate more comorbidities and thus have more conditions listed on their death certificates [12]. Conditions that predominantly kill elderly people could therefore have inflated ratios as an artefact of age. A condition like senility (R54) or falls (W00–W19) may have a high ratio partly because its decedents are frail and old, not solely because the condition is “hidden.” Our sensitivity analysis found that only 10.9% of ratio variation was explained by estimated median age at death, and no top-ranked conditions were identified as primarily age-driven, suggesting the ratio *ranking* is robust to age confounding. External validation using US CDC data showed that while age standardisation materially changed absolute ratio values for 6 of 8 cause groups (divergence 16–34%), the rank ordering was nearly perfectly preserved (Spearman ρ = 0.976 across eight cause groups, with only one adjacent rank swap). Absolute ratio values should therefore be interpreted as approximate, but the ranking of which conditions have the highest hidden burden is robust. The external validation assumes that age-correction patterns are transferable between the US and Australia given similar disease epidemiology and age structures; Australian-specific validation would require age-stratified Cube 10 data that the ABS does not currently publish. Future releases with age-specific counts would enable definitive age-standardised SRMU computation at the three-character level. Second, ratio data were available for a single year (2023), preventing multi-year analysis and limiting the power of confirmatory tests designed for paired year-level observations. Third, the ratio does not distinguish between Part I (causal chain) and Part II (contributing conditions) mentions on the death certificate, which carry different causal implications [6]. This means some mentions counted toward the ratio may be “bystander” comorbidities (conditions listed for completeness but not causally contributing to the death), making the ratio an upper bound on the true hidden burden for prevalent chronic conditions (see Discussion). Fourth, the avoidability classification may not capture the full spectrum of preventable mortality, particularly for conditions not included in the AIHW framework [17]. Finally, the publicly available ABS data do not include Indigenous status, precluding Indigenous-specific analysis. The striking mortality profile of the Northern Territory (Supplementary Figure S8), a jurisdiction where Indigenous Australians represent approximately 27% of the population [18], suggests that Indigenous health disparities are an important driver of geographic variation, but this could not be examined directly.

## Conclusions

The hidden burden of mortality in Australia is substantial: the condition at the median ratio has 60% of its death certificate mentions invisible to underlying cause statistics, and the most extreme conditions have over 99% hidden. The multiple-to-underlying ratio provides a transparent, reproducible framework for identifying conditions whose population health impact is systematically understated. While pre-registered hypotheses about sex differences did not survive multiple testing correction, the consistent pattern of higher male ratios, particularly for mental health conditions, warrants replication with multi-year data. This approach can be applied to any country publishing both underlying and multiple cause of death statistics, enabling international comparisons of hidden mortality burden.

## Data Availability

The study used ONLY openly available human data that were originally located in the following locations.
1. Australian Bureau of Statistics (ABS) - Causes of Death, Australia, 2023
Cat. No. 3303.0 (released 2024)
- Cube 10: Multiple causes of death by ICD-10 code and sex
- Cube 14: Underlying cause by year of occurrence (2014-2024)
URL: https://www.abs.gov.au/statistics/health/causes-death/causes-death-australia/latest-release
2. Australian Bureau of Statistics (ABS) - National, state and territory population
Cat. No. 3101.0, June 2023
- Estimated Resident Population by state/territory and sex
URL: https://www.abs.gov.au/statistics/people/population/national-state-and-territory-population/latest-release
3. Australian Institute of Health and Welfare (AIHW) - National Healthcare Agreement:
PI 16 - Potentially avoidable deaths, 2023
- Avoidability classification (preventable vs treatable)
URL: https://www.aihw.gov.au/reports-data/indicators/national-healthcare-agreement
4. Australian Institute of Health and Welfare (AIHW) - Australia's health 2024
- National health indicators and Indigenous health data
URL: https://www.aihw.gov.au/reports-data/australias-health
5. Australian Institute of Health and Welfare (AIHW) - Australian Burden of Disease Study 2023
- Burden of disease estimates (reference year 2018)
URL: https://www.aihw.gov.au/reports/burden-of-disease/australian-burden-of-disease-study-2023
6. US Centers for Disease Control and Prevention (CDC) - WONDER databases
- Multiple Cause of Death (D77) and Underlying Cause of Death (D76)
- Data year 2020 (used for age-standardisation validation)
URL: https://wonder.cdc.gov/
ANALYSIS CODE AND DERIVED DATA
GitHub repository (available upon publication): https://github.com/hayden-farquhar/aus-mortality-hidden-burden
Pre-registration: Open Science Framework (OSF): https://doi.org/10.17605/OSF.IO/K46RN
Note: All primary data sources are publicly available. Raw ABS Excel files and AIHW data were downloaded from the URLs above. The analysis code and all derived datasets (CSVs, figures) are available in the GitHub repository.

https://github.com/hayden-farquhar/aus-mortality-hidden-burden

## Data availability

All data used in this study are publicly available from the Australian Bureau of Statistics (Causes of Death 2023) [16] and the Australian Institute of Health and Welfare (burden of disease [14], avoidable deaths classification [17]). Analysis code and derived data are available at https://github.com/hayden-farquhar/aus-mortality-hidden-burden.

## Acknowledgements

The author gratefully acknowledges the Australian Bureau of Statistics for the provision of Causes of Death data and population estimates, and the Australian Institute of Health and Welfare for the National Healthcare Agreement avoidable deaths classification and burden of disease data. This study would not have been possible without the sustained investment in high-quality, publicly accessible health data by these agencies.

## Author contributions

HF conceived and designed the study, performed all analyses, and wrote the manuscript. HF is the sole author and guarantor.

## Ethics statement

No ethics approval was required. All data analysed in this study are publicly released by the Australian Bureau of Statistics and the Australian Institute of Health and Welfare in aggregated, de-identified form. No individual-level or restricted-access data were used.

