## Supplementary for "The Hidden Burden of Mortality Across the Spectrum of ICD-10 Conditions in Australia: A Multiple Cause of Death Analysis"

Hayden Farquhar, MBBS MPHTM

February 2026

### Supplementary Note: Structure of the Australian death certificate

Australian death certificates follow the WHO International Form of Medical Certificate of Cause of Death [1]. The certificate has two parts:

- **Part I** records the causal sequence leading to death, from the immediate cause (line a) back to the underlying cause (the lowest completed line). For example: (a) Pulmonary embolism, due to (b) Immobilisation following hip fracture, due to (c) Fall at home, where "Fall at home" is selected as the underlying cause.
- **Part II** records other significant conditions that contributed to death but were not part of the direct causal sequence. For example: Type 2 diabetes mellitus, Hypertension, Chronic kidney disease.

All conditions in both Part I and Part II are counted as "multiple cause" mentions. Only the single condition identified as the underlying cause (the lowest line in Part I) enters standard mortality tabulations. The ratio captures how often a condition appears anywhere on the certificate (Parts I and II combined) relative to how often it is selected as the underlying cause. A high ratio indicates a condition that frequently contributes to death but is rarely identified as having initiated the fatal sequence.

---

### Supplementary Table S1. Complete ratio ranking for conditions with ≥10 underlying cause deaths (top 50 of 663 shown)

The full table for all 663 conditions is available as a CSV file (`mur_ranking_full.csv`) in the data repository. Conditions are ranked by descending ratio (persons). Hidden deaths = multiple cause mentions − underlying cause deaths. Sex ratio = male ratio / female ratio.

| Rank | ICD-10 | Condition | Underlying | Multiple | Ratio | % Hidden | Male Ratio | Female Ratio | Sex ratio |
|---|---|---|---|---|---|---|---|---|---|
| 1 | Y83–Y84 | Surgical and medical procedure complications | 15 | 4,217 | 281.1 | 99.6 | 475.6 | 183.9 | 2.59 |
| 2 | R57 | Shock, NEC | 24 | 3,442 | 143.4 | 99.3 | 178.0 | 114.2 | 1.56 |
| 3 | R10–R19 | Digestive symptoms and signs | 20 | 2,403 | 120.2 | 99.2 | 185.3 | 85.1 | 2.18 |
| 4 | F40–F48 | Neurotic, stress-related and somatoform disorders | 22 | 2,325 | 105.7 | 99.1 | 230.0 | 69.1 | 3.33 |
| 5 | R64 | Cachexia | 14 | 1,382 | 98.7 | 99.0 | 183.7 | 75.5 | 2.43 |
| 6 | Y40–Y84 | Complications of medical and surgical care | 68 | 6,413 | 94.3 | 98.9 | 106.6 | 82.1 | 1.30 |
| 7 | F41 | Other anxiety disorders | 19 | 1,739 | 91.5 | 98.9 | 269.3 | 58.2 | 4.63 |
| 8 | J96 | Respiratory failure, NEC | 92 | 7,582 | 82.4 | 98.8 | 87.3 | 77.9 | 1.12 |

| Rank | ICD-10 | Condition | Underlying | Multiple | Ratio | % Hidden | Male Ratio | Female Ratio | Sex ratio |
|---|---|---|---|---|---|---|---|---|---|
| 9 | R40–R46 | Cognitive, perceptual and emotional symptoms and signs | 55 | 4,250 | 77.3 | 98.7 | 139.0 | 49.7 | 2.80 |
| 10 | R63 | Food and fluid intake symptoms and signs | 23 | 1,746 | 75.9 | 98.7 | 90.3 | 69.6 | 1.30 |
| 11 | F15 | Mental and behavioural disorders: other stimulants | 16 | 1,063 | 66.4 | 98.5 | 70.5 | 54.2 | 1.30 |
| 12 | F32 | Depressive episode | 71 | 3,723 | 52.4 | 98.1 | 81.2 | 37.8 | 2.15 |
| 13 | R00–R09 | Circulatory and respiratory symptoms and signs | 75 | 3,876 | 51.7 | 98.1 | 67.1 | 41.4 | 1.62 |
| 14 | Y40–Y59 | Drugs and biologicals causing adverse effects | 42 | 2,121 | 50.5 | 98.0 | 50.3 | 50.7 | 0.99 |
| 15 | F30–F39 | Mood [affective] disorders | 92 | 4,306 | 46.8 | 97.9 | 72.3 | 33.9 | 2.14 |
| 16 | R41 | Cognitive function symptoms and signs | 55 | 2,433 | 44.2 | 97.7 | 63.8 | 35.5 | 1.80 |

| Rank | ICD-10 | Condition | Underlying | Multiple | Ratio | % Hidden | Male Ratio | Female Ratio | Sex ratio |
|---|---|---|---|---|---|---|---|---|---|
| 17 | R09 | Other circulatory and respiratory symptoms and signs | 63 | 2,545 | 40.4 | 97.5 | 49.2 | 34.2 | 1.44 |
| 18 | I95–I99 | Other circulatory system disorders | 23 | 916 | 39.8 | 97.5 | 40.2 | 39.4 | 1.02 |
| 19 | F05 | Delirium (non-substance) | 62 | 2,449 | 39.5 | 97.5 | 65.7 | 27.9 | 2.36 |
| 20 | M50–M54 | Other dorsopathies | 15 | 592 | 39.5 | 97.5 | 84.0 | 23.3 | 3.61 |
| 21 | G45 | Transient cerebral ischaemic attacks | 26 | 955 | 36.7 | 97.3 | 39.7 | 34.9 | 1.14 |
| 22 | I99 | Other and unspecified circulatory system disorders | 15 | 531 | 35.4 | 97.2 | 35.2 | 35.6 | 0.99 |
| 23 | R53 | Malaise and fatigue | 329 | 10,347 | 31.4 | 96.8 | 40.1 | 27.5 | 1.46 |
| 24 | J81 | Pulmonary oedema | 60 | 1,796 | 29.9 | 96.7 | 30.8 | 29.2 | 1.06 |

| Rank | ICD-10 | Condition | Underlying | Multiple | Ratio | % Hidden | Male Ratio | Female Ratio | Sex ratio |
|---|---|---|---|---|---|---|---|---|---|
| 25 | F19 | Mental and behavioural disorders: multiple drug use | 23 | 686 | 29.8 | 96.6 | 32.0 | 24.9 | 1.29 |
| 26 | J95–J99 | Other respiratory system diseases | 337 | 9,830 | 29.2 | 96.6 | 32.1 | 26.6 | 1.21 |
| 27 | I10 | Essential (primary) hypertension | 844 | 23,420 | 27.7 | 96.4 | 41.8 | 21.2 | 1.98 |
| 28 | F31 | Bipolar affective disorder | 20 | 543 | 27.1 | 96.3 | 45.8 | 19.1 | 2.39 |
| 29 | M10 | Gout | 24 | 628 | 26.2 | 96.2 | 34.6 | 14.3 | 2.42 |
| 30 | M19 | Other arthrosis | 98 | 2,351 | 24.0 | 95.8 | 39.2 | 20.1 | 1.95 |
| 31 | G47 | Sleep disorders | 92 | 2,177 | 23.7 | 95.8 | 24.9 | 21.8 | 1.14 |
| 32 | M81 | Osteoporosis without pathological fracture | 100 | 2,326 | 23.3 | 95.7 | 28.1 | 22.2 | 1.26 |
| 33 | R50–R69 | General symptoms and signs | 1,478 | 32,073 | 21.7 | 95.4 | 29.8 | 18.0 | 1.65 |
| 34 | D70 | Agranulocytosis | 20 | 433 | 21.6 | 95.4 | 34.4 | 14.8 | 2.33 |
| 35 | F20–F29 | Schizophrenia and delusional disorders | 69 | 1,482 | 21.5 | 95.3 | 29.0 | 15.3 | 1.90 |
| 36 | F20 | Schizophrenia | 55 | 1,127 | 20.5 | 95.1 | 25.7 | 15.5 | 1.66 |

| Rank | ICD-10 | Condition | Underlying | Multiple | Ratio | % Hidden | Male Ratio | Female Ratio | Sex ratio |
|---|---|---|---|---|---|---|---|---|---|
| 37 | F10–F19 | Substance use disorders | 503 | 10,466 | 20.8 | 95.2 | 19.7 | 24.9 | 0.79 |
| 38 | E87 | Other fluid, electrolyte and acid-base disorders | 93 | 1,929 | 20.7 | 95.2 | 25.0 | 18.4 | 1.36 |
| 39 | N17 | Acute renal failure | 333 | 6,755 | 20.3 | 95.1 | 21.2 | 19.2 | 1.10 |
| 40 | R50 | Fever of other or unknown origin | 16 | 320 | 20.0 | 95.0 | 24.0 | 17.6 | 1.36 |
| 41 | E86 | Volume depletion | 122 | 2,329 | 19.1 | 94.8 | 28.1 | 14.6 | 1.92 |
| 42 | N19 | Unspecified renal failure | 164 | 3,075 | 18.8 | 94.7 | 19.9 | 17.6 | 1.13 |
| 43 | N17–N19 | Renal failure | 498 | 9,060 | 18.2 | 94.5 | 18.9 | 17.3 | 1.09 |
| 44 | D65–D69 | Coagulation defects and haemorrhagic conditions | 45 | 805 | 17.9 | 94.4 | 20.1 | 16.0 | 1.26 |
| 45 | R54 | Senility | 508 | 9,028 | 17.8 | 94.4 | 19.5 | 16.9 | 1.15 |
| 46 | D69 | Purpura and other haemorrhagic conditions | 38 | 666 | 17.5 | 94.3 | 20.3 | 15.3 | 1.33 |
| 47 | G90–G99 | Other nervous system disorders | 115 | 1,956 | 17.0 | 94.1 | 25.5 | 12.2 | 2.09 |
| 48 | G93 | Other brain disorders | 90 | 1,448 | 16.1 | 93.8 | 22.1 | 12.8 | 1.73 |

| Rank | ICD-10 | Condition | Underlying | Multiple | Ratio | % Hidden | Male Ratio | Female Ratio | Sex ratio |
|---|---|---|---|---|---|---|---|---|---|
| 49 | H60–H95 | Ear and mastoid process diseases | 19 | 287 | 15.1 | 93.4 | 15.8 | 14.3 | 1.10 |
| 50 | E10–E14 | Diabetes mellitus | 4,510 | 61,430 | 13.6 | 92.7 | 14.2 | 13.0 | 1.09 |

*Note: The full ranking of 663 conditions is available as Supplementary Data File (`mur_ranking_full.csv`). Distributional summary: median ratio = 2.5, mean = 6.6; 71 conditions (10.7%) had ratio > 10; 267 (40.3%) had ratio 2.0–10; 325 (49.0%) had ratio < 2.0. NEC = not elsewhere classified.* ***Notable exception:*** *Rank 37, Substance use disorders (F10–F19), is the only major block where the female ratio exceeds male (sex ratio = 0.79), counter to the general male-excess pattern.* ***Interpretive caveat:*** *The ratio treats all certificate mentions equally, both causally relevant contributions and incidental comorbidities ("bystander" mentions). It therefore provides an upper bound on the true hidden burden, particularly for highly prevalent chronic conditions (e.g., hypertension) that certifiers may list routinely regardless of causal role in the specific death. Some high-ratio conditions (e.g., respiratory failure J96, ratio = 82.4) reflect WHO coding rules that classify them as "modes of dying" rather than initiating causes, a convention-driven rather than clinically-driven high ratio. Sex-specific ratios use sex-specific numerators and denominators.*

### Supplementary Table S2. Sex-divergent conditions: male/female ratio > 2.0 or < 0.5

Conditions where the male ratio is more than double the female ratio (sex ratio > 2.0) or less than half (sex ratio < 0.5), indicating substantial sex differences in the relative hidden mortality burden. Sorted by descending sex ratio. Inclusion criteria: ≥10 underlying cause deaths (persons) for the main ratio ranking; the sex ratio column additionally requires ≥10 underlying deaths in each sex separately (conditions where either sex had zero underlying deaths are excluded as the sex ratio is undefined).

| ICD-10 | Condition | Ratio (persons) | Male Ratio | Female Ratio | Sex ratio | Underlying deaths |
|---|---|---|---|---|---|---|
| K90 | Intestinal malabsorption | 5.2 | 37.0 | 6.5 | 5.73 | 14 |
| F41 | Other anxiety disorders | 91.5 | 269.3 | 58.2 | 4.63 | 19 |
| K29 | Gastritis and duodenitis | 5.1 | 12.0 | 2.7 | 4.50 | 36 |
| E51 | Thiamine deficiency | 4.1 | 8.0 | 1.8 | 4.36 | 18 |
| M50–M54 | Other dorsopathies | 39.5 | 84.0 | 23.3 | 3.61 | 15 |
| F40–F48 | Neurotic, stress-related and somatoform disorders | 105.7 | 230.0 | 69.1 | 3.33 | 22 |
| R40–R46 | Cognitive, perceptual and emotional symptoms | 77.3 | 139.0 | 49.7 | 2.80 | 55 |
| Y83–Y84 | Surgical and medical procedure complications | 281.1 | 475.6 | 183.9 | 2.59 | 15 |
| R64 | Cachexia | 98.7 | 183.7 | 75.5 | 2.43 | 14 |
| M10 | Gout | 26.2 | 34.6 | 14.3 | 2.42 | 24 |

| ICD-10 | Condition | Ratio (persons) | Male Ratio | Female Ratio | Sex ratio | Underlying deaths |
|---|---|---|---|---|---|---|
| F31 | Bipolar affective disorder | 27.1 | 45.8 | 19.1 | 2.39 | 20 |
| F05 | Delirium (non-substance) | 39.5 | 65.7 | 27.9 | 2.36 | 62 |
| D70 | Agranulocytosis | 21.6 | 34.4 | 14.8 | 2.33 | 20 |
| R10–R19 | Digestive symptoms and signs | 120.2 | 185.3 | 85.1 | 2.18 | 20 |
| F32 | Depressive episode | 52.4 | 81.2 | 37.8 | 2.15 | 71 |
| F30–F39 | Mood [affective] disorders | 46.8 | 72.3 | 33.9 | 2.14 | 92 |
| G90–G99 | Other nervous system disorders | 17.0 | 25.5 | 12.2 | 2.09 | 115 |
| M05–M14 | Inflammatory polyarthropathies | 7.1 | 10.9 | 5.5 | 2.00 | 93 |

*Note: 28 conditions had male/female ratio > 2.0 (shown: those with ≥10 underlying deaths in both sexes). Only one condition had sex ratio < 0.5: acute tubulo-interstitial nephritis (N10; sex ratio = 0.47, male ratio = 1.5, female ratio = 3.2). Conditions where either sex had zero underlying deaths are excluded (sex ratio undefined). Sex ratio > 1 indicates higher relative hidden burden in males.*

### Supplementary Table S3. Holm-Bonferroni multiple testing correction

All eight statistical tests from the three confirmatory hypotheses, ranked by ascending uncorrected p-value, with Holm-Bonferroni corrections applied across primary tests (3 tests) and across all tests (8 tests). No test achieved significance after correction.

| Holm Rank | Test ID | Hypothesis | Test description | Type | p (uncorrected) | p (Holm, primary) | p (Holm, all) | Significant? |
|---|---|---|---|---|---|---|---|---|
| 1 | H1b | H1 | Hypertension sex gap temporal trend (Spearman, crude rates) | Secondary | 0.0146 | - | 0.116 | No |
| 2 | H3a | H3 | Mental health ratio: male vs female (Wilcoxon signed-rank) | Primary | 0.0234 | 0.070 | 0.164 | No |
| 3 | H3b_nonsub | H3 | Non-substance (F20–F99) ratio: male vs female (Wilcoxon) | Secondary | 0.0313 | - | 0.188 | No |
| 4 | H3c | H3 | Mental health sex gap temporal trend (Spearman, crude rates) | Secondary | 0.0388 | - | 0.194 | No |
| 5 | H1a | H1 | Hypertension ratio: male vs female (Wilcoxon signed-rank) | Primary | 0.125 | 0.250 | 0.500 | No |
| 6 | H3b_sub | H3 | Substance use (F10–F19) ratio: male vs female (Wilcoxon) | Secondary | 0.625 | - | 1.000 | No |

| Holm Rank | Test ID | Hypothesis | Test description | Type | p (uncorrected) | p (Holm, primary) | p (Holm, all) | Significant? |
|---|---|---|---|---|---|---|---|---|
| 7 | H2b | H2 | Geographic CV: preventable vs treatable (Mann-Whitney U) | Secondary | 0.832 | - | 1.000 | No |
| 8 | H2a | H2 | Geographic CV: avoidable vs non-avoidable (Mann-Whitney U) | Primary | 0.872 | 0.872 | 1.000 | No |

*Note: Holm-Bonferroni correction was applied in two tiers: (1) across 3 primary tests (one per hypothesis), and (2) across all 8 tests. Secondary tests were conditional on the primary test achieving significance. The "-" entries for secondary test p (Holm, primary) indicate these tests were not included in the primary-level correction. The closest to significance was H3a (mental health ratio sex difference): uncorrected p = 0.023, but p_holm = 0.070 after primary-level correction.*

---

### Supplementary Table S4. Confirmatory hypothesis test results with sensitivity analyses

#### S4a. H1: Hypertension sex-differentiated hidden burden

| Test | Statistic | Value | p (uncorrected) | Effect size | n | Note |
|---|---|---|---|---|---|---|
| Wilcoxon signed-rank (primary) | Z | 1.83 | 0.125 | r = 0.77 | 4 | Adapted from n=17 years to n=4 sub-conditions |
| Exact binomial sign test | n positive | 4 | 0.125 | - | 4 | All 4 sub-conditions showed male > female ratio |
| Bootstrap CI (BCa) | Mean diff | 5.49 | - | CI: [0.24, 15.71] | 4 | 95% CI excludes zero |
| Permutation test | Mean diff | 5.49 | 0.124 | - | 4 | 10,000 permutations |

**Input data (H1):**

| Condition | ICD-10 | Male Ratio | Female Ratio | Difference | Direction |
|---|---|---|---|---|---|
| Essential hypertension | I10 | 41.8 | 21.2 | +20.6 | Male > Female |
| Hypertensive heart disease | I11 | 2.3 | 1.5 | +0.8 | Male > Female |
| Hypertensive renal disease | I12 | 1.4 | 1.2 | +0.2 | Male > Female |
| Hypertensive heart and renal disease | I13 | 1.9 | 1.7 | +0.2 | Male > Female |

*Note: Ratios and differences are displayed rounded to 1 decimal place. The mean difference of 5.49 and bootstrap CI [0.24, 15.71] were computed from full-precision values (e.g., I10 male = 41.833, female = 21.160, diff = 20.673), which is why the mean of the displayed rounded differences (5.45) differs slightly. The Wilcoxon Z statistic is the standardised test statistic from the coin package (wilcoxsign_test with exact distribution), not the traditional sum-of-ranks V.*

#### S4b. H2: Geographic variation by preventability

| Test | Statistic | Value | p (uncorrected) | Effect size | n (groups) | Note |
|---|---|---|---|---|---|---|
| Mann-Whitney U (primary) | Z | −0.16 | 0.872 | r_rb = 0.012 | 100 vs 151 | Avoidable vs non-avoidable |
| Permutation test | Median diff | −0.30 | 0.906 | - | 100 vs 151 | 10,000 permutations |
| Mann-Whitney U (secondary) | U | 1,135 | 0.832 | r_rb = −0.027 | 67 vs 33 | Preventable vs treatable |
| Sensitivity (excl. NT and ACT) | U | 7,459 | 0.872 | r_rb = 0.012 | 100 vs 151 | Small-population jurisdictions excluded |

**Summary statistics:** Median CV for avoidable conditions = 30.8%; non-avoidable = 31.1%. Preventable = 30.1%; treatable = 32.6%.

#### S4c. H3: Mental health sex-differentiated hidden burden

| Test | Statistic | Value | p (uncorrected) | Effect size | n | Note |
|---|---|---|---|---|---|---|
| Wilcoxon signed-rank (primary) | Z | 2.24 | 0.023 | r = 0.80 | 8 | Adapted from n=17 years to n=8 block-level sub-conditions |
| Exact binomial sign test | n positive | 7 | 0.070 | - | 8 | 7 of 8 blocks showed male > female ratio |
| Bootstrap CI (percentile) | Mean diff | 40.8 | - | CI: [7.69, 82.33] | 8 | 95% CI excludes zero |
| Permutation test | Mean diff | 40.8 | 0.025 | - | 8 | 10,000 permutations |
| Non-substance sub-analysis | Z | 2.20 | 0.031 | r = 0.88 | 6 | F20–F99: 6 block-level sub-conditions |
| Substance use sub-analysis | Z | 0.67 | 0.625 | r = 0.22 | 5 | F10–F19: 5 three-character codes with ≥10 underlying deaths in both sexes |

**Input data (H3, block-level):**

| Block | ICD-10 | Male Ratio | Female Ratio | Difference | Direction |
|---|---|---|---|---|---|
| Organic and symptomatic mental disorders | F00–F09 | 2.6 | 2.2 | +0.4 | Male > Female |
| Substance use disorders | F10–F19 | 19.7 | 24.9 | −5.2 | Female > Male |
| Schizophrenia and delusional disorders | F20–F29 | 29.0 | 15.3 | +13.7 | Male > Female |
| Mood disorders | F30–F39 | 72.3 | 33.9 | +38.4 | Male > Female |
| Neurotic and stress-related disorders | F40–F48 | 230.0 | 69.1 | +160.9 | Male > Female |
| Behavioural and physiological syndromes | F50–F59 | 19.3 | 6.7 | +12.7 | Male > Female |
| Intellectual disability | F70–F79 | 37.3 | 30.4 | +6.8 | Male > Female |
| Developmental disorders | F80–F89 | 120.0 | 21.5 | +98.5 | Male > Female |

*Note: Eight blocks with valid male and female ratios were used as paired observations. Three blocks were excluded: F60–F69 (Personality disorders; 0 female underlying deaths, female ratio undefined), F90–F98 (Childhood behavioural disorders; 0 underlying deaths for both sexes), and F99 (Unspecified; 0 underlying deaths). The only included block where the female ratio exceeded the male ratio was substance use disorders (F10–F19). This was contrary to the pre-registered prediction that substance use would drive the sex difference. Instead, non-substance disorders showed the stronger sex effect ($r = 0.88$ vs $r = 0.22$).*

### Supplementary Table S5. Suicide-driven ratio inflation analysis

Counterfactual estimates quantifying the contribution of suicide coding rules to the male mental health ratio excess. When a death is classified as suicide, the underlying cause is assigned to External Causes (X60–X84) per WHO coding rules [1], while any contributing mental health conditions are recorded only as multiple causes. This mechanically inflates the ratio for mental health codes.

**Central estimate (50% mention rate):**

| Metric | Male | Female | Sex Ratio |
|---|---|---|---|
| Observed ratio | 4.92 | 3.04 | 1.62 |
| Counterfactual ratio (suicide mentions removed) | 4.67 | 2.99 | 1.56 |
| Difference explained | 0.25 | 0.05 | 0.06 |
| % of excess explained | - | - | 11% |

**Sensitivity analysis:**

| Assumed mention rate | Male ratio (counterfactual) | Female ratio (counterfactual) | Sex ratio (counterfactual) | % of excess explained |
|---|---|---|---|---|
| 30% | 4.77 | 3.01 | 1.58 | 6% |
| 50% (central) | 4.67 | 2.99 | 1.56 | 11% |
| 70% | 4.56 | 2.96 | 1.54 | 15% |
| 90% (extrapolated) | ~4.46 | ~2.94 | ~1.52 | ~19% |
| 100% (theoretical max) | ~4.41 | ~2.93 | ~1.50 | ~21% |

*Note: The 50% central estimate is based on US surveillance data showing 46% of suicide decedents had a known mental health condition [20]. The 90% and 100% rows are linear extrapolations from the 30–70% pattern (~2 percentage points per 10% increase in mention rate); Australian coronial practices may support mention rates toward the upper end of this range, but even at the theoretical maximum the suicide coding mechanism explains only approximately one-fifth of the male ratio excess.*

### Supplementary Table S6. Age-adjustment sensitivity analysis

Raw versus age-adjusted ratio rankings for conditions with ≥50 underlying deaths. Full table available as CSV file (`age_adjusted_mur_ranking.csv`). Note: ABS Cube 10 provides aggregated condition-level counts without age stratification; unit record files that would enable direct age-standardisation at the three-character ICD-10 level are restricted-access microdata. The indirect and regression-based adjustments below use chapter-level median age estimates as proxies.

**Summary findings:** - Regression $R^2 = 0.109$: Only 10.9% of ratio variation explained by estimated median age at death - No conditions identified as primarily "age-driven" (top 30 raw → below rank 50 after adjustment) - Top 15 high-ratio conditions retained approximate rankings after both adjustment methods

### Supplementary Table S7. CDC WONDER age-standardisation validation results

**Data source:** US CDC WONDER Multiple Cause of Death (D77) and Underlying Cause of Death (D76) databases, 2020. Direct age standardisation using US 2000 Standard Population (Census P25-1130) across 11 ten-year age groups.

| Cause group | ICD-10 codes | Crude Ratio | Age-standardised Ratio | Difference | Material (>10%)? |
|---|---|---|---|---|---|
| Diabetes mellitus | E10–E14 | 3.80 | 2.49 | −34.4% | Yes |
| Alzheimer disease | G30 | 1.30 | 1.57 | +20.7% | Yes |
| Hypertensive diseases | I10, I12, I15 | 12.84 | 10.33 | −19.6% | Yes |
| Ischaemic heart disease | I20–I25 | 1.61 | 1.73 | +7.1% | No |
| Cerebrovascular disease | I60–I69 | 1.83 | 2.14 | +17.3% | Yes |
| Influenza and pneumonia | J09–J18 | 5.98 | 4.98 | −16.7% | Yes |
| Chronic lower respiratory diseases | J40–J47 | 2.36 | 2.37 | +0.6% | No |
| Renal failure | N17–N19 | 4.71 | 5.45 | +15.6% | Yes |

**Summary:** Of 8 cause groups, 6 showed material divergence (>10%) between crude and age-standardised ratios, while 2 were concordant (ischaemic heart disease: +7.1%; CLRD: +0.6%). For diabetes, hypertension, and influenza/pneumonia (conditions concentrated in the elderly), age standardisation reduced the ratio by 17–34%, suggesting the crude ratio overestimates hidden burden for these conditions. Conversely, cerebrovascular disease, renal failure, and Alzheimer disease showed 16–21% higher age-standardised ratios.

*Note: Despite the material divergence in absolute values, the rank ordering was nearly perfectly preserved: the Spearman rank correlation between crude and age-standardised rankings across the eight cause groups was $\rho = 0.976$, with only one adjacent rank swap (influenza/pneumonia and renal failure exchanging positions 2 and 3). This validation uses US data as a proxy for the age-correction effect, assuming transferability between the US and Australia given similar disease epidemiology and age structures. The full validation data are available as* `cdc_mur_validation_results.csv` *in the repository.*

### Supplementary Figures

**Supplementary Figure S1. Absolute hidden burden by ICD-10 chapter**

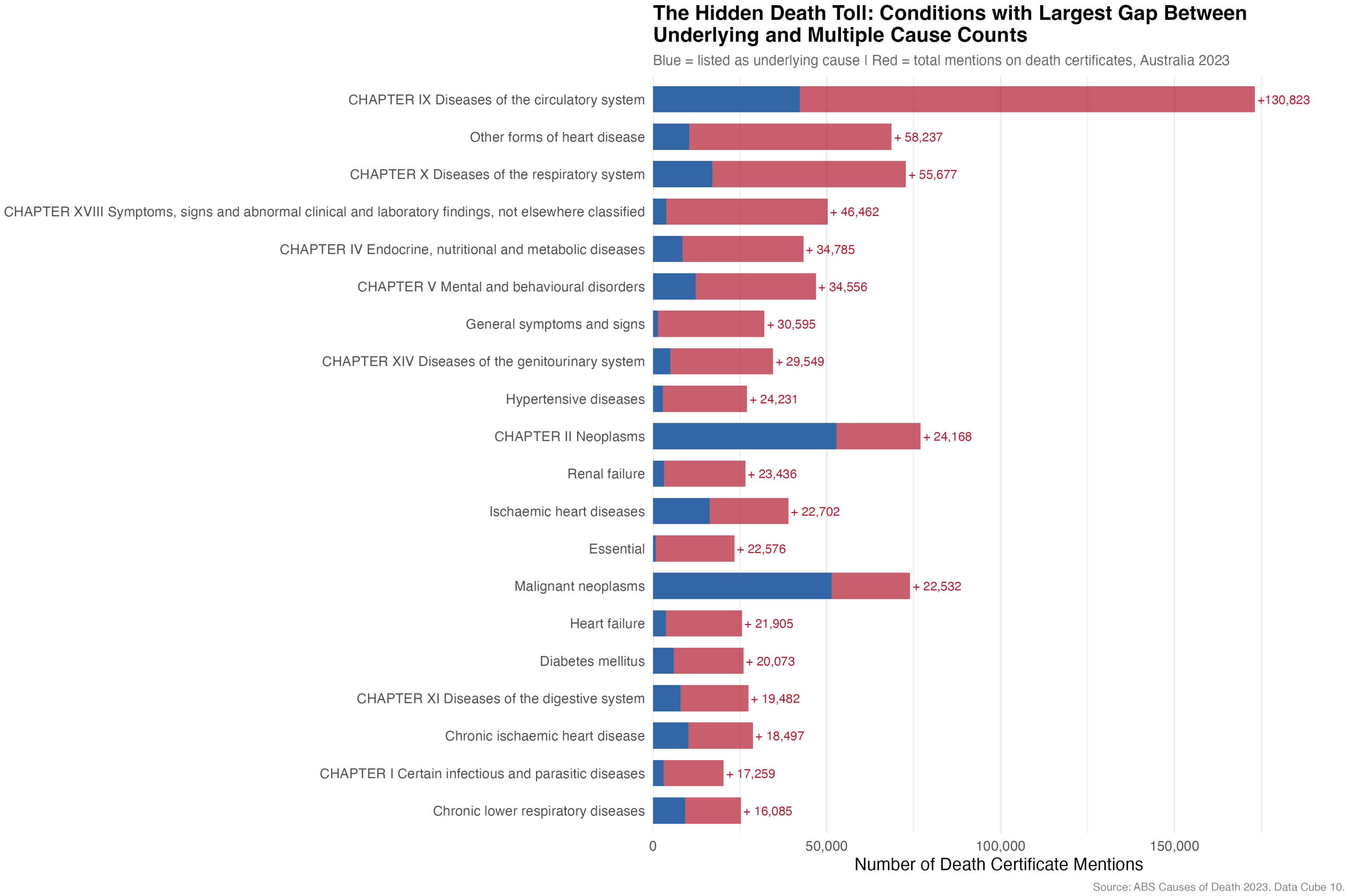

Supplementary Figure S1

**Supplementary Figure S1.** Absolute number of hidden death certificate mentions by ICD-10 chapter (multiple cause mentions minus underlying cause deaths). The circulatory system chapter has the largest absolute hidden burden (130,823 hidden mentions), despite having a moderate ratio of 4.1, due to the very large number of deaths in this chapter.

**Supplementary Figure S2. Hidden burden ratio by ICD-10 chapter**

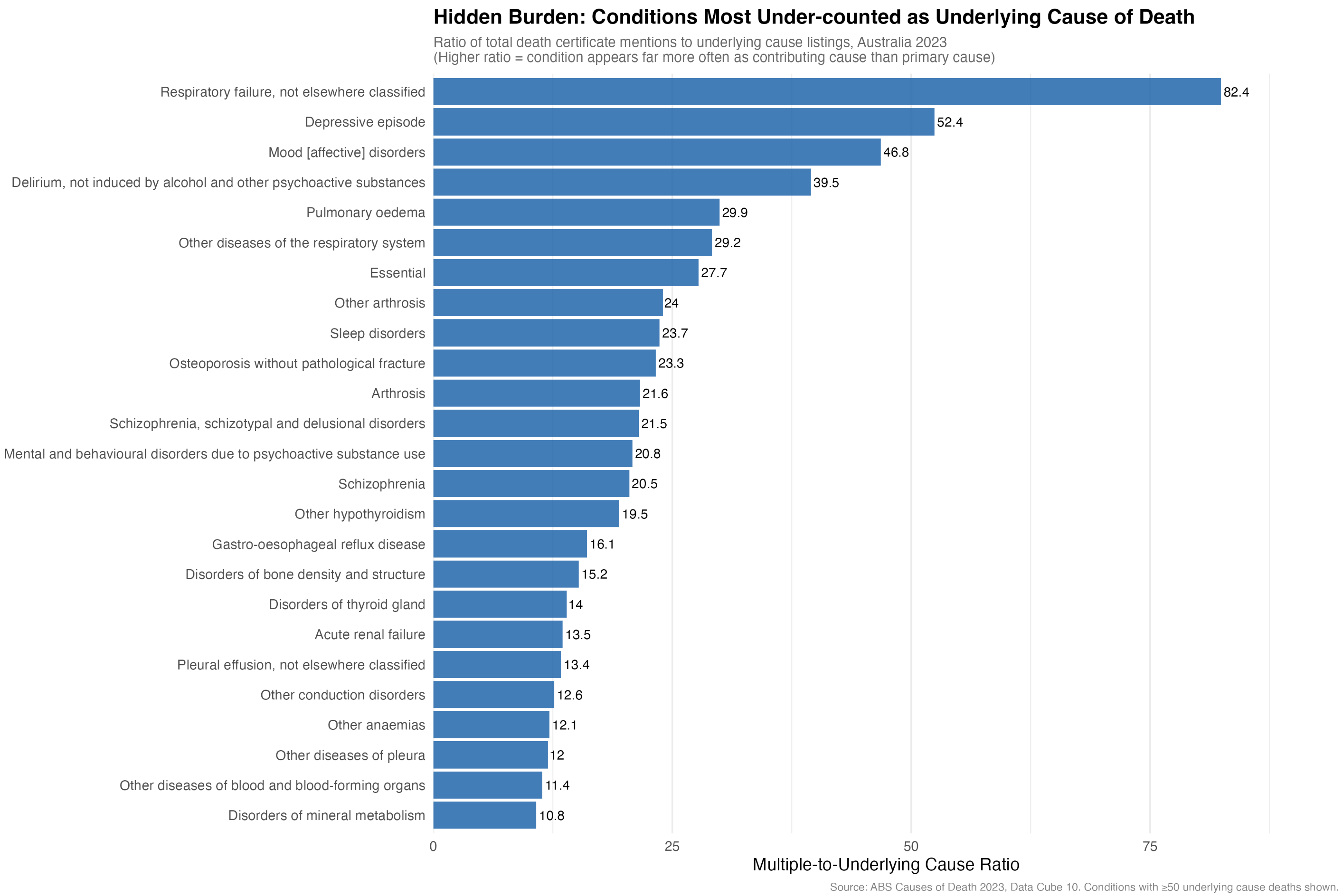

Supplementary Figure S2

**Supplementary Figure S2.** Hidden burden ratio by ICD-10 chapter, showing the proportion of death certificate mentions hidden from underlying cause statistics. This figure was moved from the main text to focus the main manuscript on code-level rather than chapter-level findings.

**Supplementary Figure S3. Conditions most frequently reported as the sole cause of death**

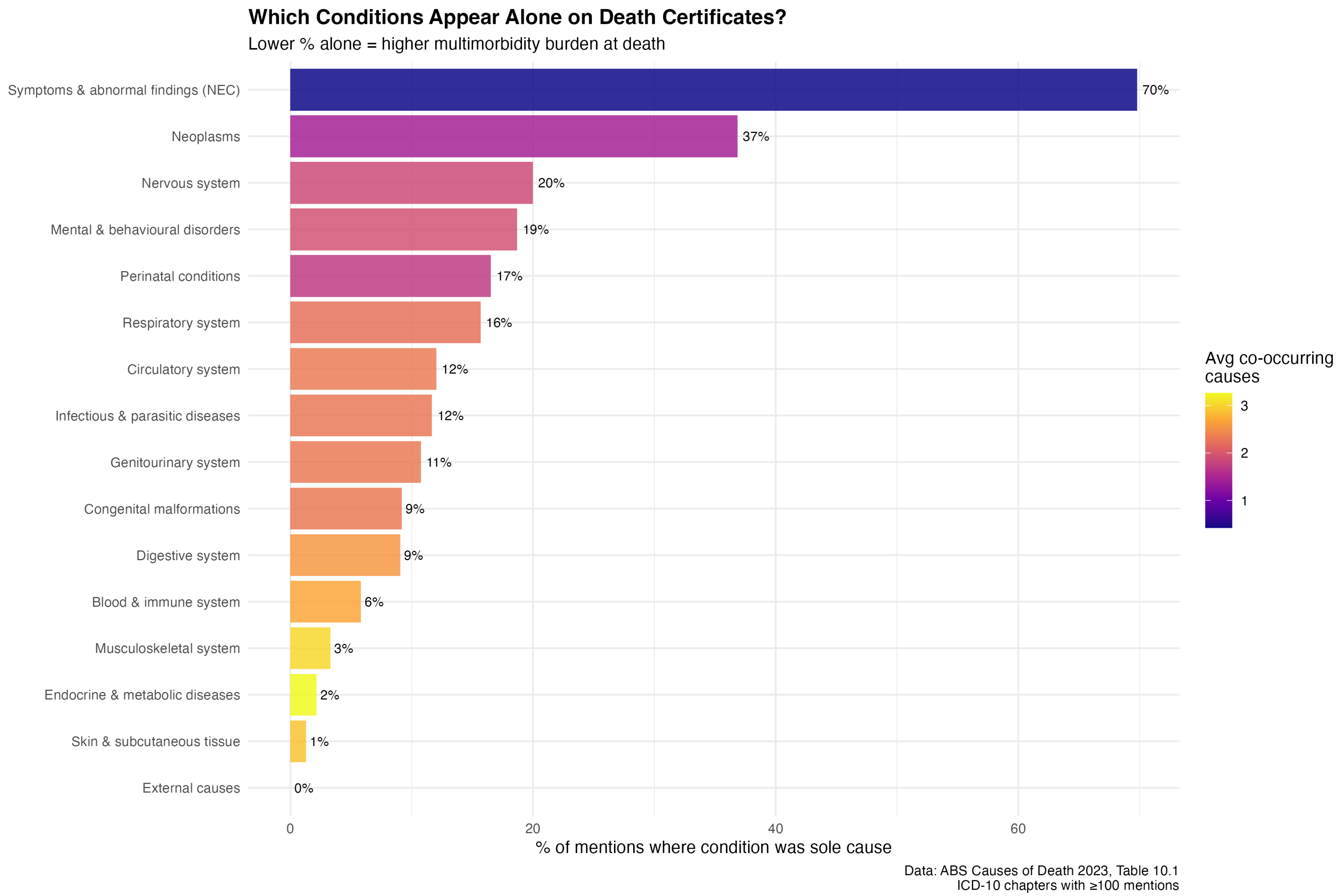

Supplementary Figure S3

**Supplementary Figure S3.** Percentage of death certificates where each condition was reported as the sole (only) cause of death. External causes (suicide, accidents, assault) are most commonly the sole cause, consistent with their low ratios. Conditions rarely reported alone are embedded in complex multi-morbidity chains.

### Supplementary Figure S4. Conditions with the highest multimorbidity

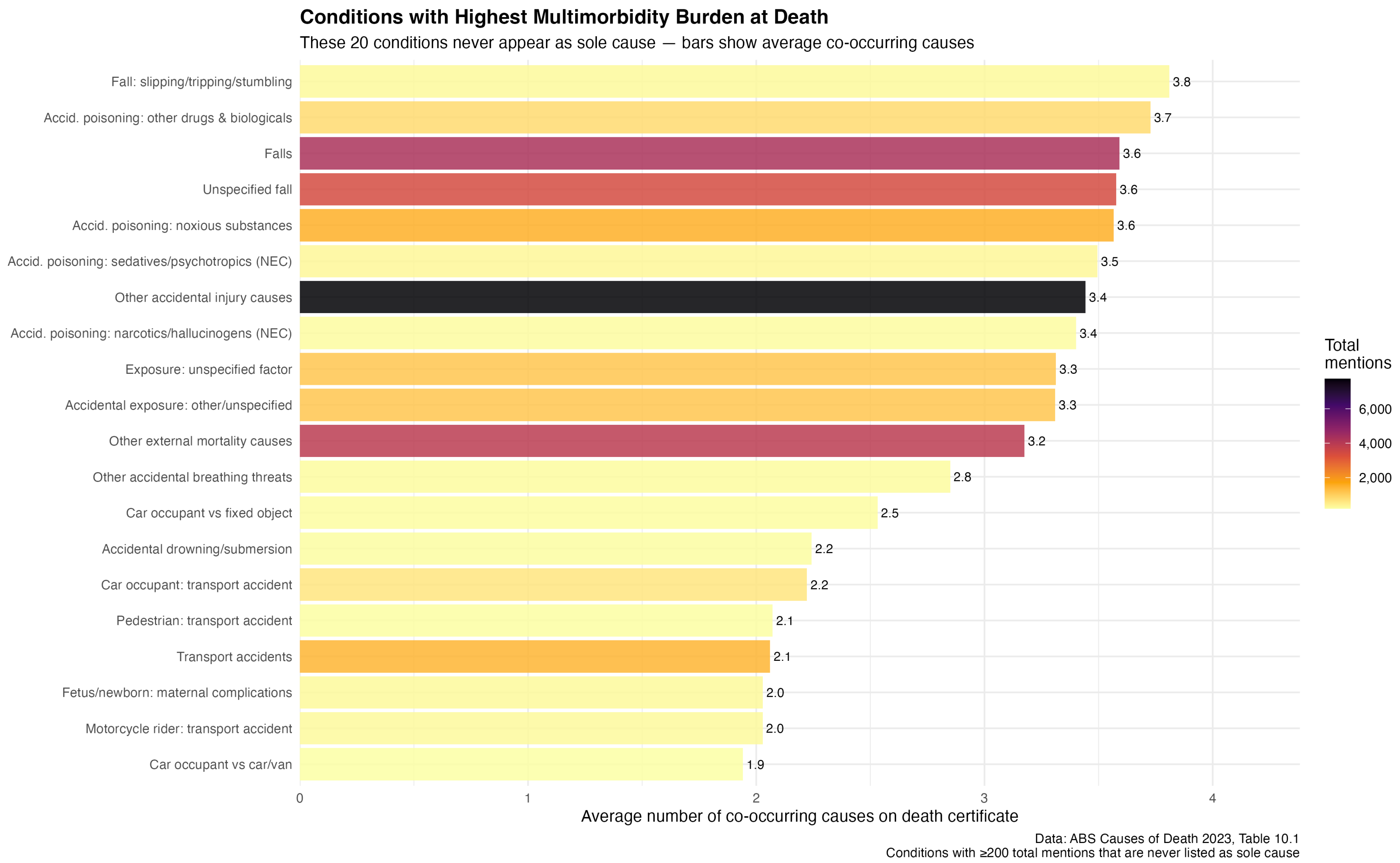

Supplementary Figure S4

**Supplementary Figure S4.** Conditions most commonly reported with three or more co-causes on the death certificate. High multimorbidity overlap indicates conditions frequently embedded in complex causal chains, consistent with high ratio values.

**Supplementary Figure S5. Sex-stratified ratio scatter plot**

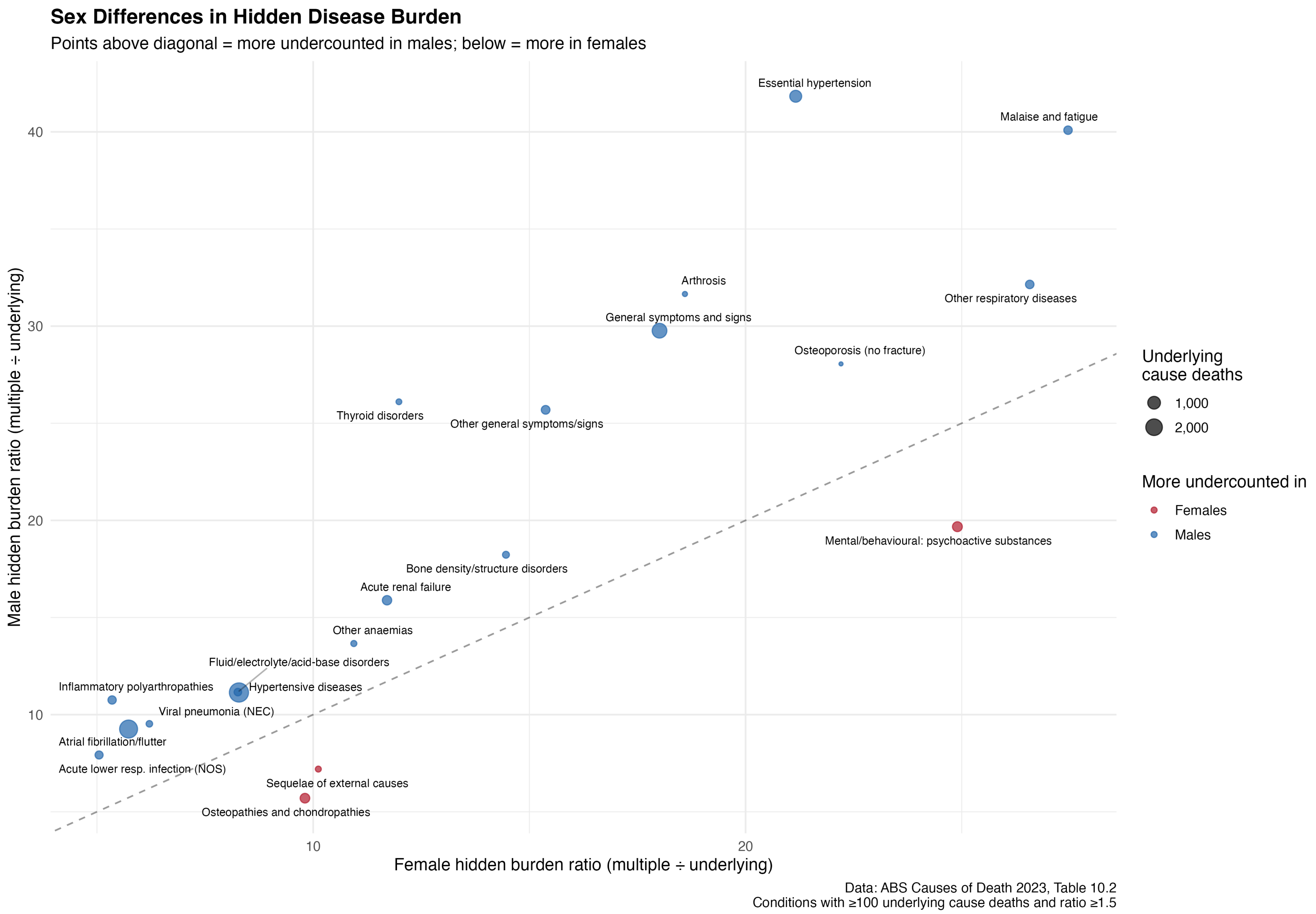

Supplementary Figure S5

**Supplementary Figure S5.** Scatter plot of male versus female ratio for all ICD-10 blocks. Points above the diagonal indicate conditions with higher male ratios (greater relative hidden burden in males). Most blocks show higher male ratios, with the exception of substance use disorders.

**Supplementary Figure S6. Cardiovascular sex-stratified hidden burden**

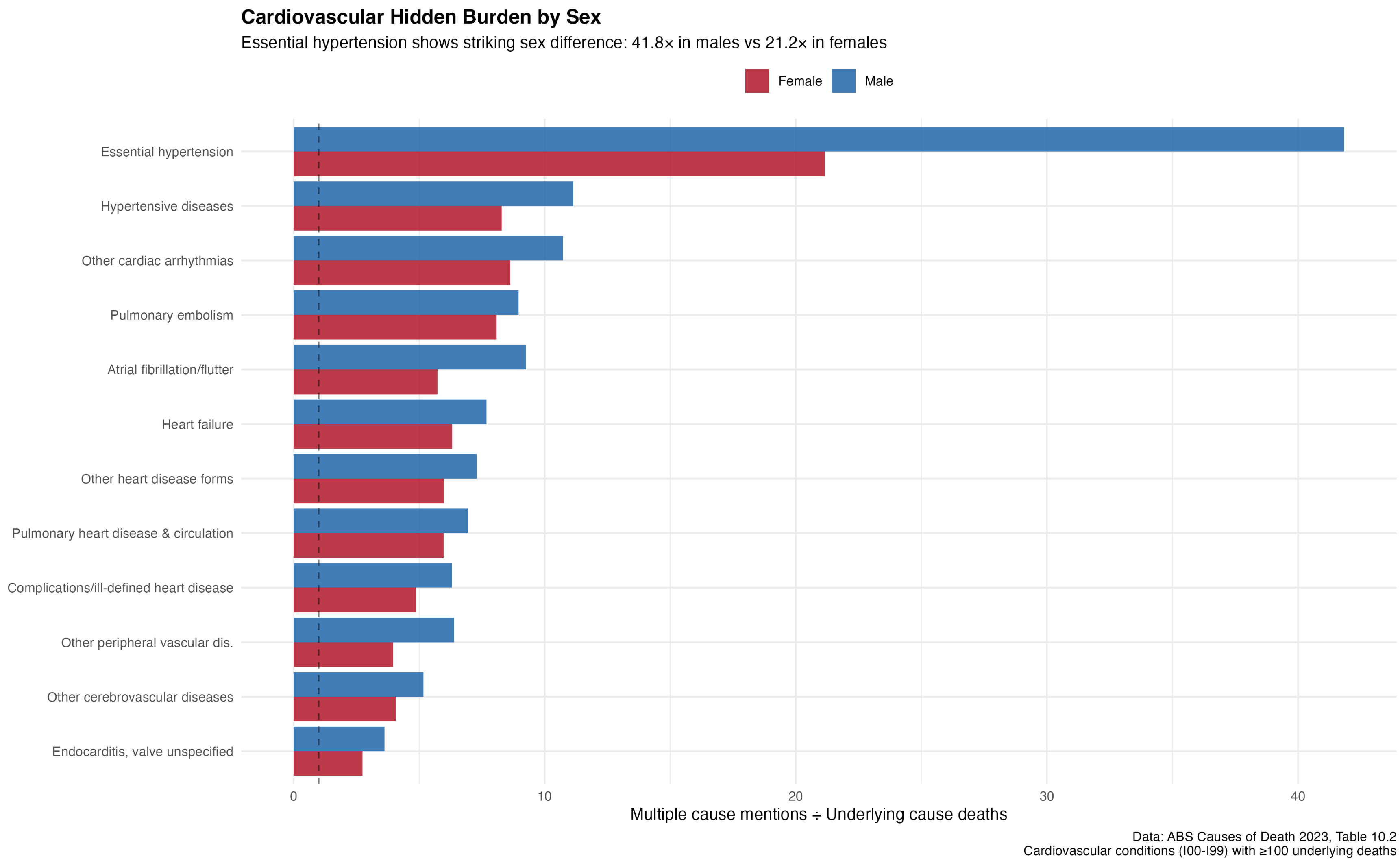

Supplementary Figure S6

**Supplementary Figure S6.** Sex-stratified ratios for cardiovascular sub-conditions (I00–I99). Essential hypertension (I10) shows the most pronounced sex difference (male ratio = 41.8, female ratio = 21.2), while most other cardiovascular conditions show relatively similar male and female ratio values.

**Supplementary Figure S7. Geographic variation in mortality rates (coefficient of variation)**

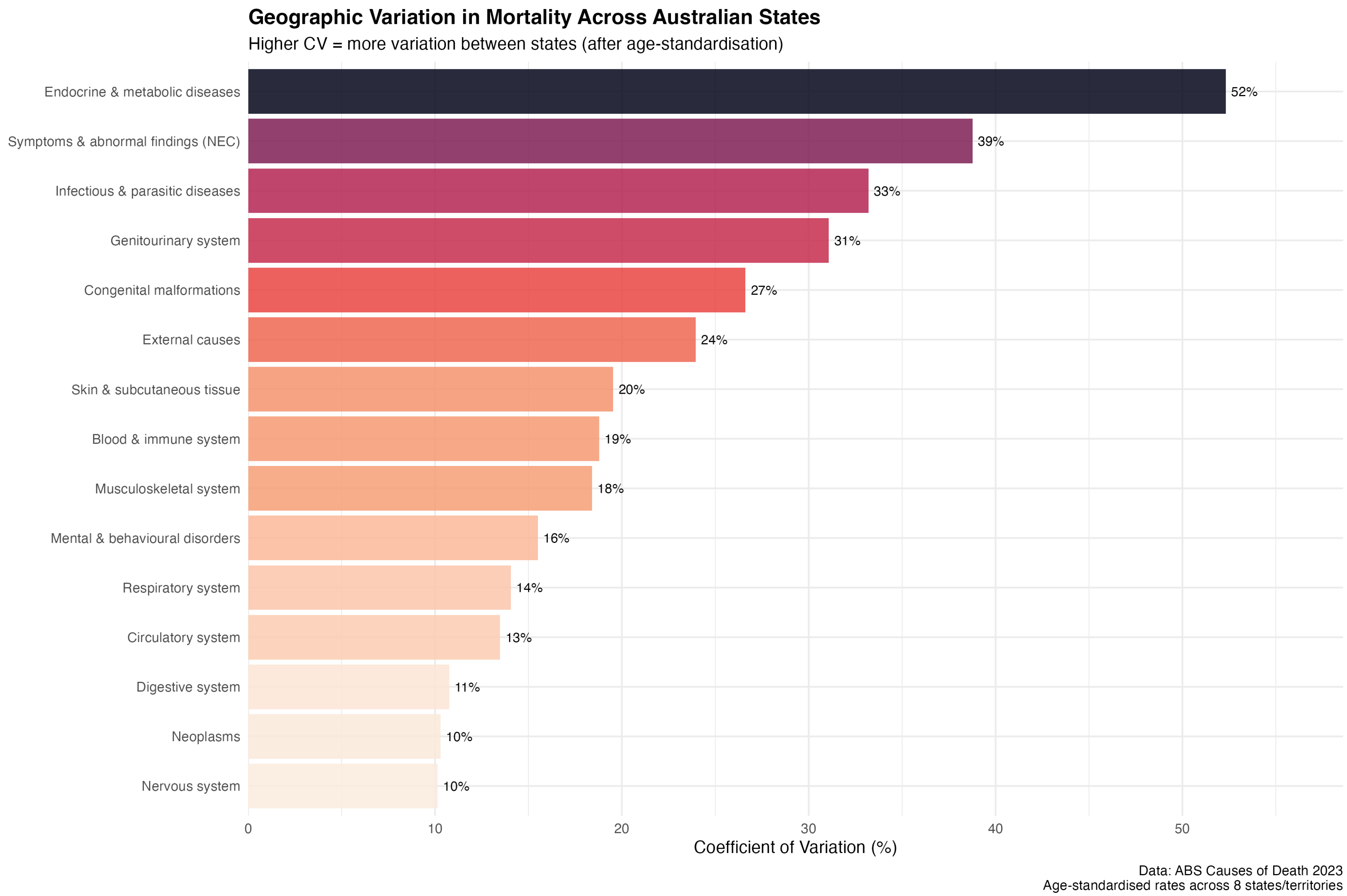

Supplementary Figure S7

**Supplementary Figure S7.** Coefficient of variation (CV) in age-standardised mortality rates across eight Australian states and territories, for all conditions with available data. The distribution of CVs is similar for avoidable and non-avoidable conditions, consistent with the null result for Hypothesis H2.

### Supplementary Figure S8. State mortality heatmap

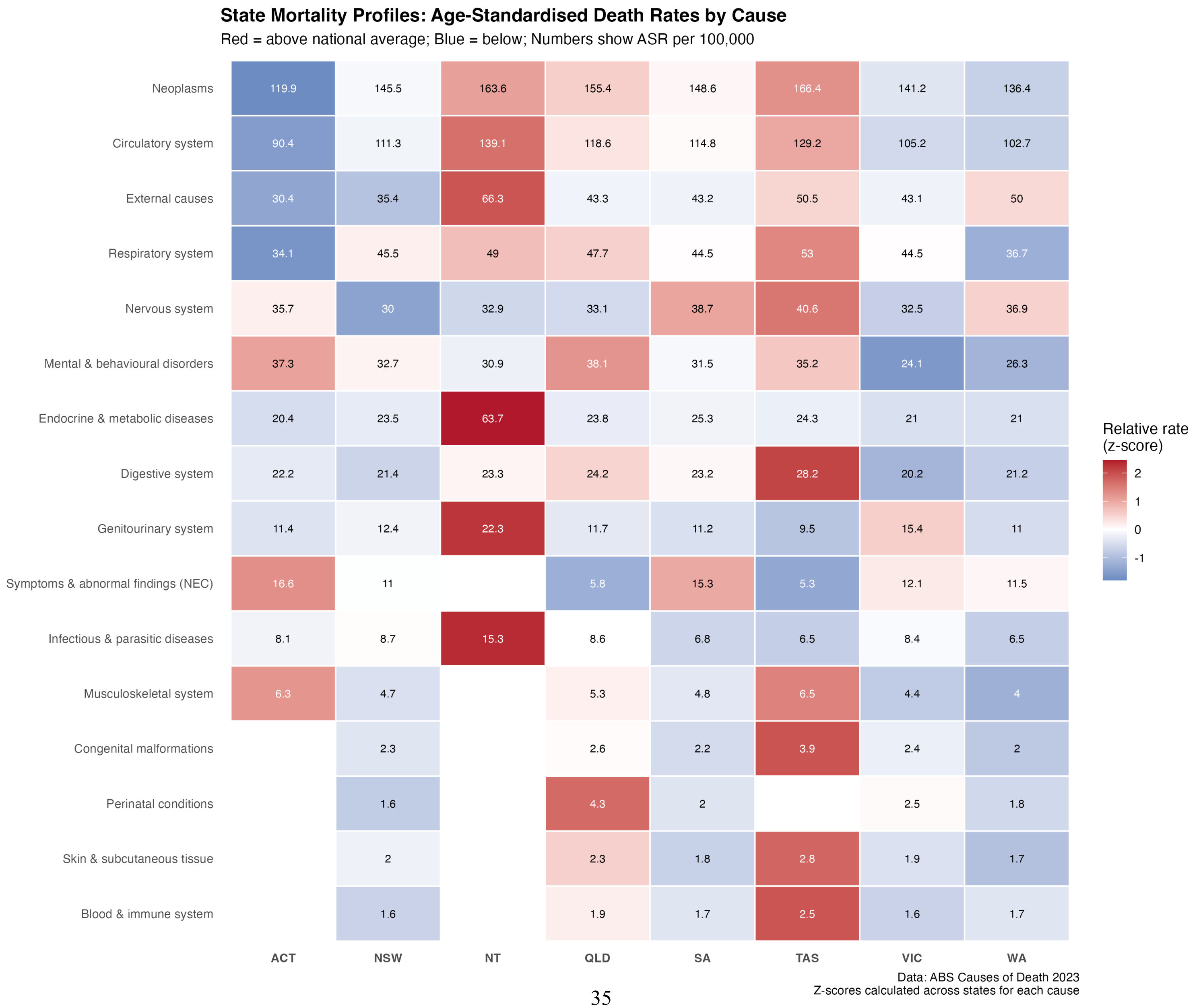

Supplementary Figure S8

**Supplementary Figure S8.** Heatmap of age-standardised mortality rates by state/territory and ICD-10 chapter. Colour intensity indicates rate magnitude. The Northern Territory shows consistently higher rates across most chapters, reflecting the well-documented health disparities affecting the Indigenous population [21].

**Supplementary Figure S9. Geographic variation for specific conditions**

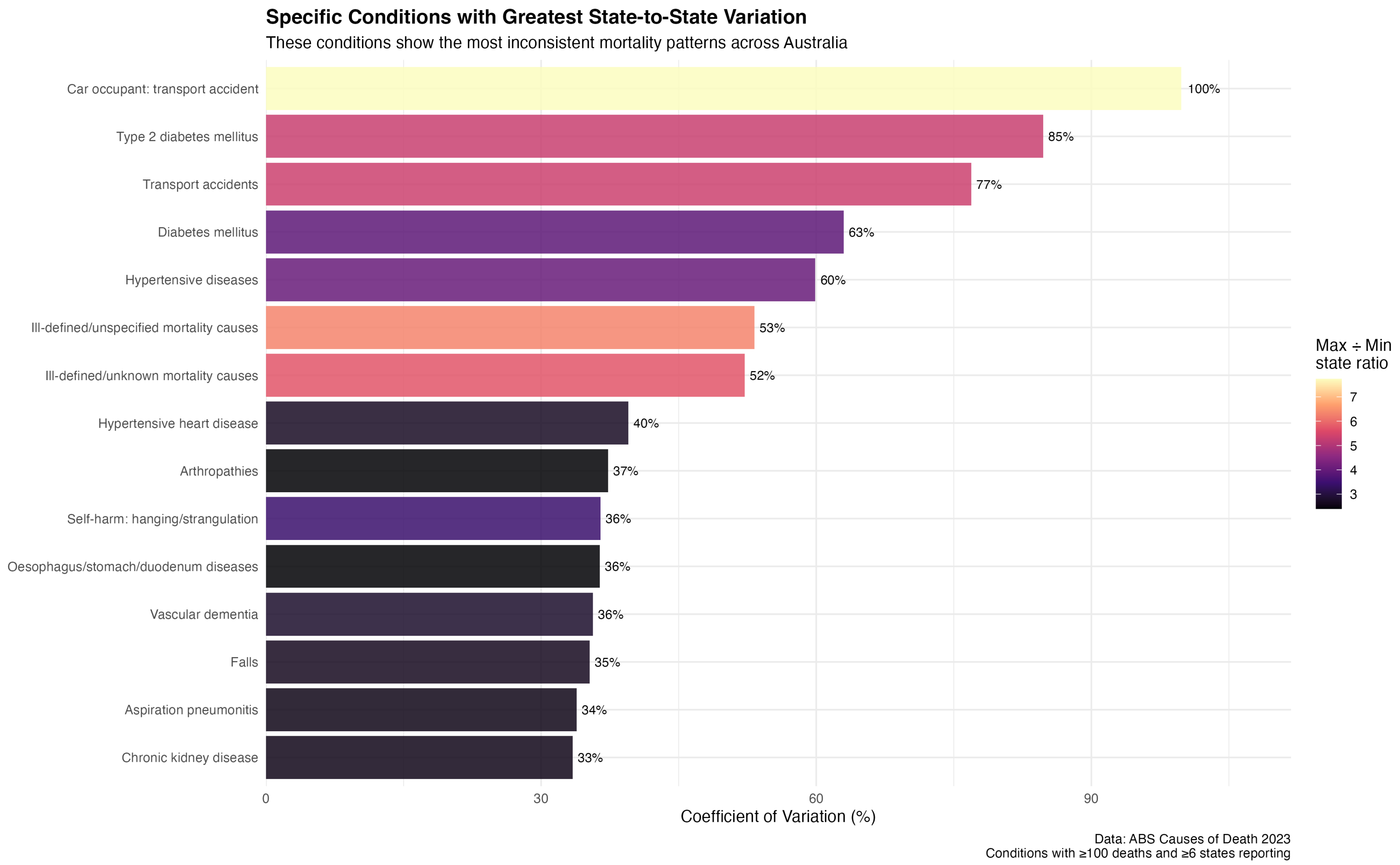

Supplementary Figure S9

**Supplementary Figure S9.** Geographic variation in mortality rates for selected high-burden conditions. Horizontal bars show the range of age-standardised rates across states and territories. Conditions with larger ranges show greater geographic inequality in mortality.

**Supplementary Figure S10. Hypothesis H1 results: Hypertension ratio by sex**

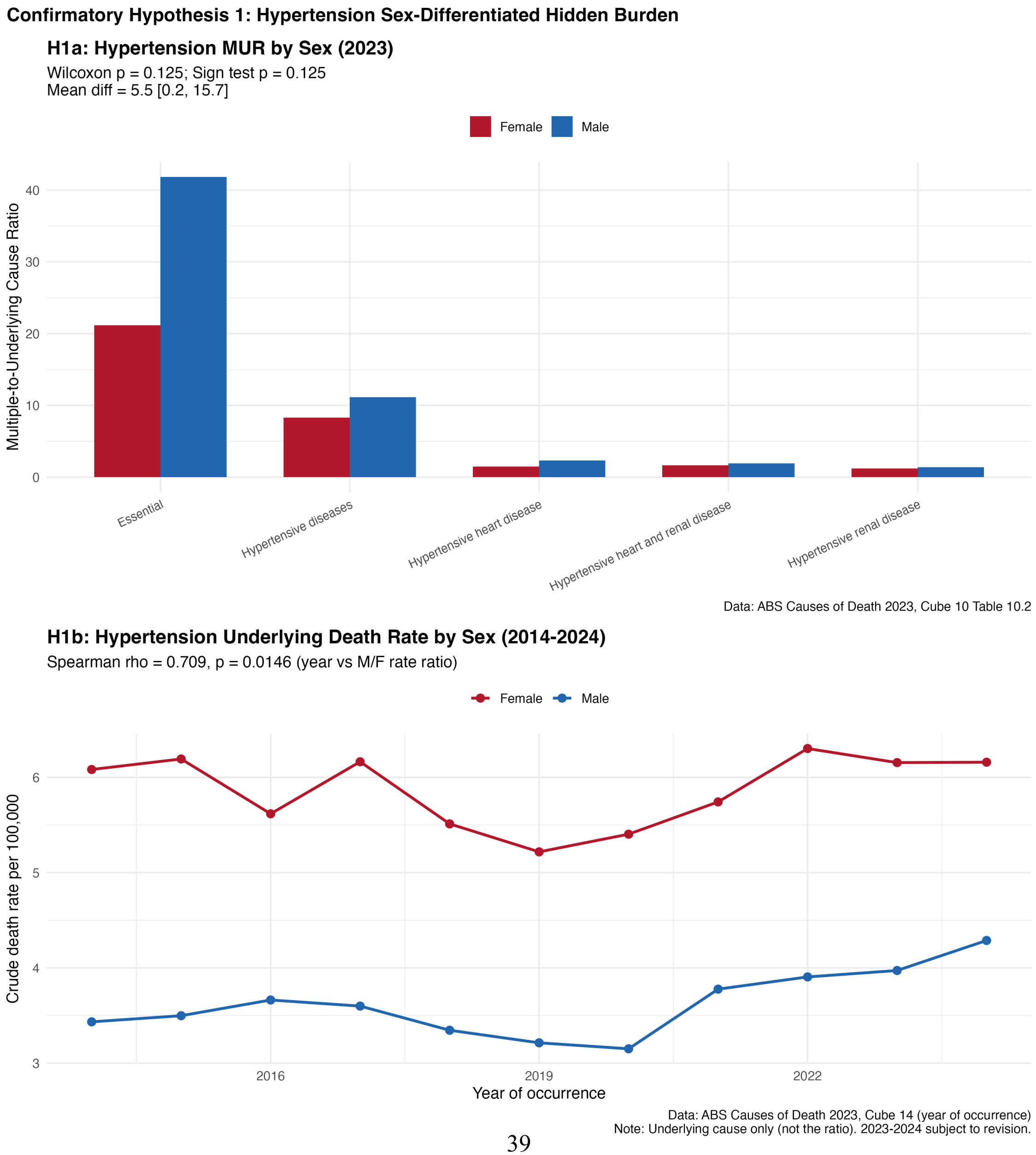

Supplementary Figure S10

**Supplementary Figure S10.** Detailed results for Hypothesis H1 (hypertension sex-differentiated hidden burden). Panel shows male versus female ratio for hypertensive disease sub-conditions (I10–I13). All four sub-conditions show higher male ratios, with essential hypertension (I10) showing the largest absolute difference. With $n = 4$ paired observations, the minimum achievable two-tailed p-value for a Wilcoxon signed-rank test is 0.125 (Holm-Bonferroni corrected $p = 0.25$), meaning the test was structurally incapable of reaching $\alpha = 0.05$. The observed $p = 0.125$ represents the strongest possible result for this sample size, not an ambiguous null.

---

**Supplementary Figure S11. Hypothesis H2 results: Geographic variation by avoidability**

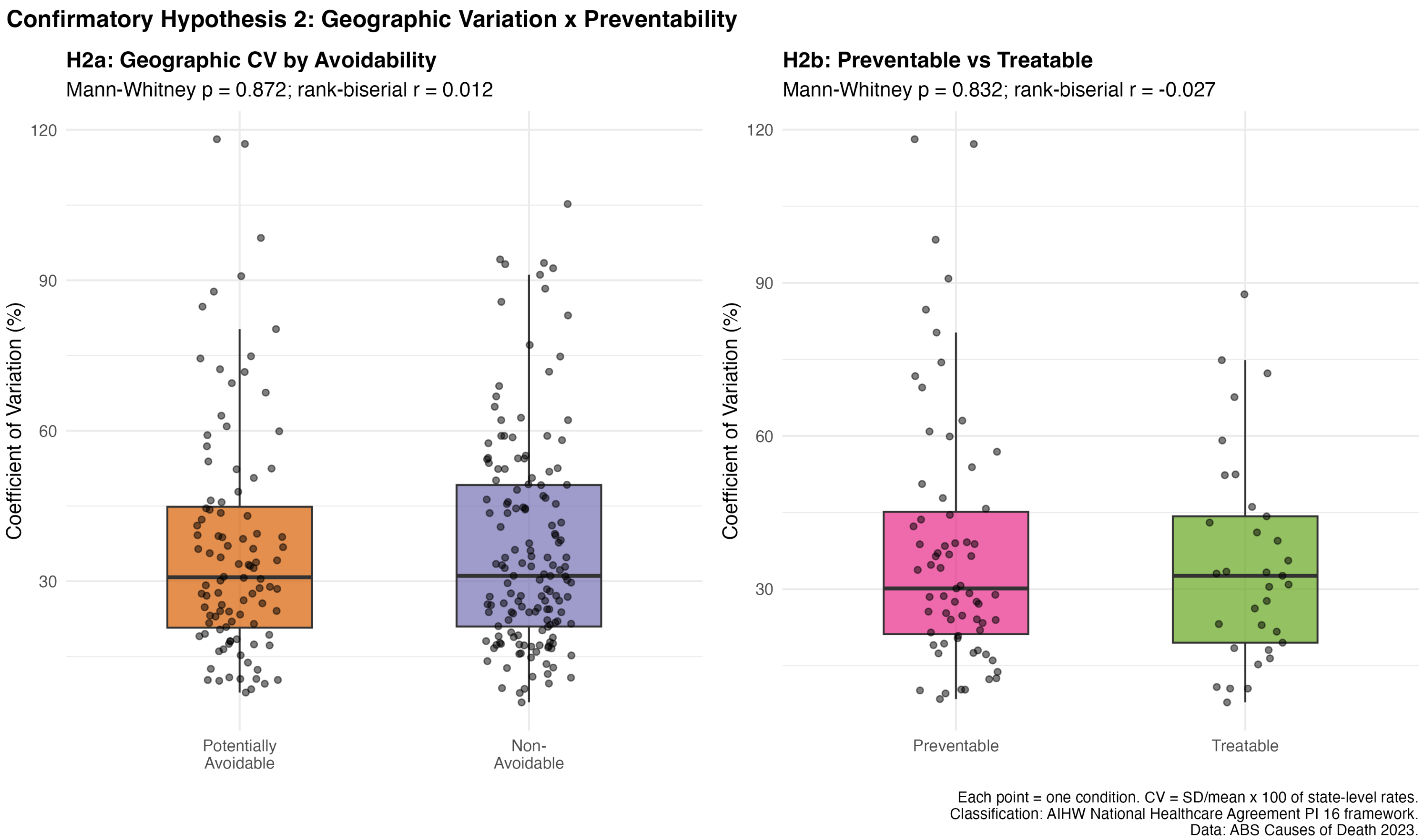

Supplementary Figure S11

**Supplementary Figure S11.** Detailed results for Hypothesis H2 (geographic variation by preventability). Distributions of coefficient of variation (CV) for avoidable versus non-avoidable conditions. The near-complete overlap confirms the clear null finding (Mann-Whitney U, $p = 0.872$).

**Supplementary Figure S12. Sex-stratified ratio ranking (top 20 conditions)**

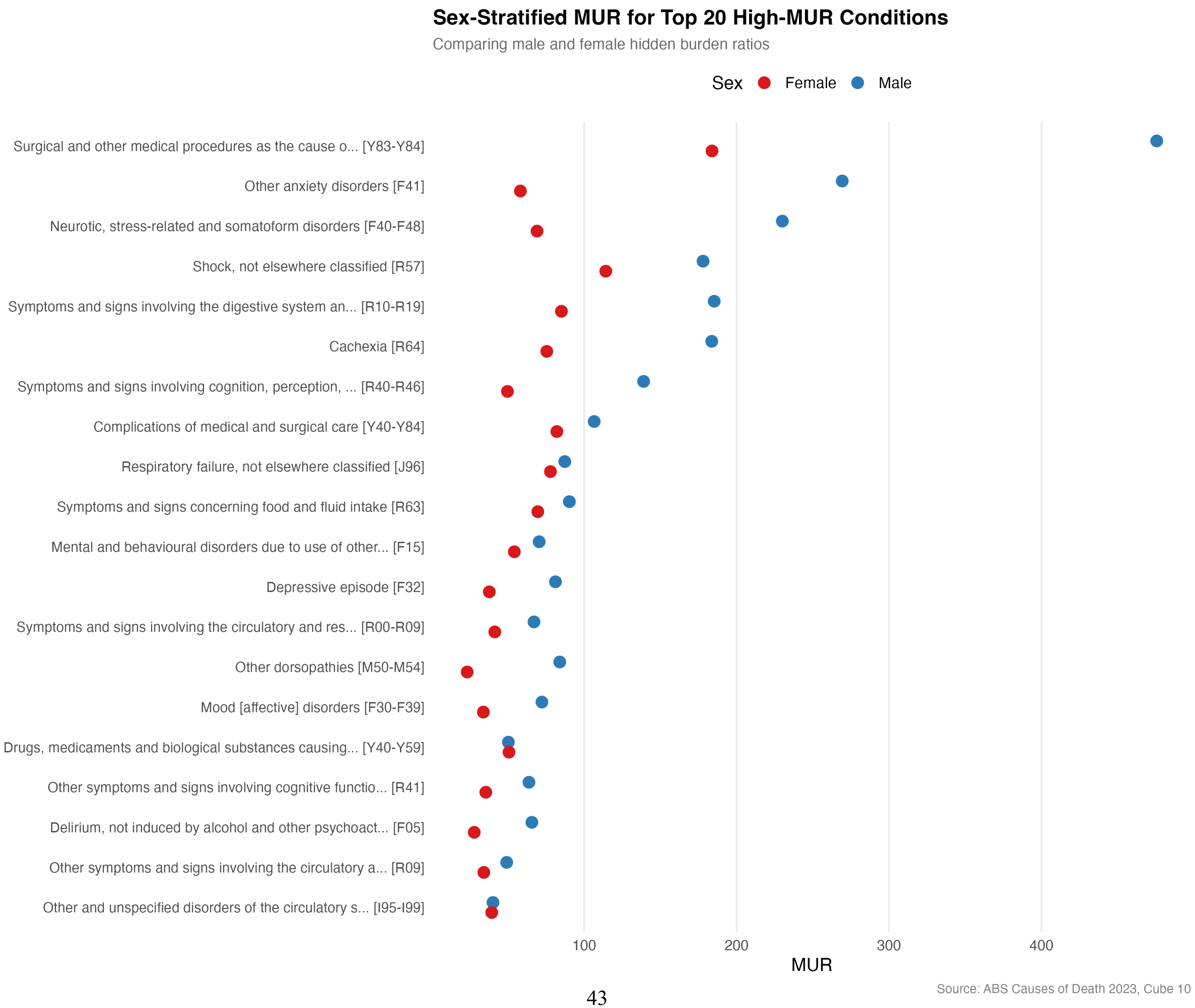

Supplementary Figure S12

**Supplementary Figure S12.** Male (blue) and female (red) ratios for the top 20 conditions ranked by persons ratio. For most conditions, the male ratio exceeds the female ratio, with the sex gap being particularly pronounced for anxiety disorders (F41), neurotic disorders (F40–F48), and cachexia (R64).

---

**Supplementary Figure S13. Age-adjustment sensitivity analysis: observed versus expected ratio**

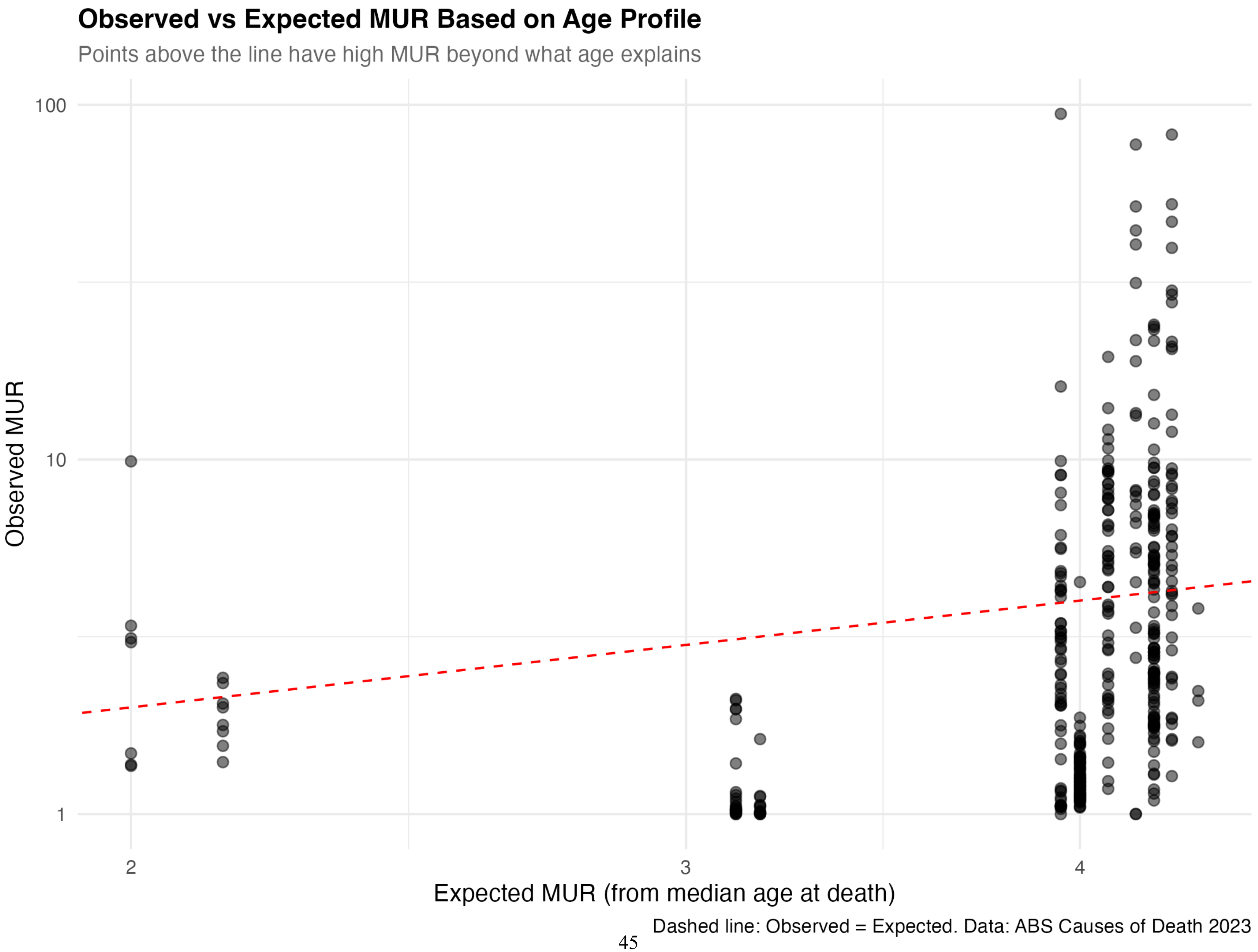

Supplementary Figure S13

**Supplementary Figure S13.** Observed ratio versus expected ratio based on estimated median age at death. Points above the diagonal have higher ratios than predicted by age alone; points below have lower ratios than age would predict. The weak relationship ($R^2 = 0.109$) indicates that age explains only a small fraction of ratio variation.

---

**Supplementary Figure S14. Age-adjustment sensitivity analysis: regression of log(ratio) on median age**

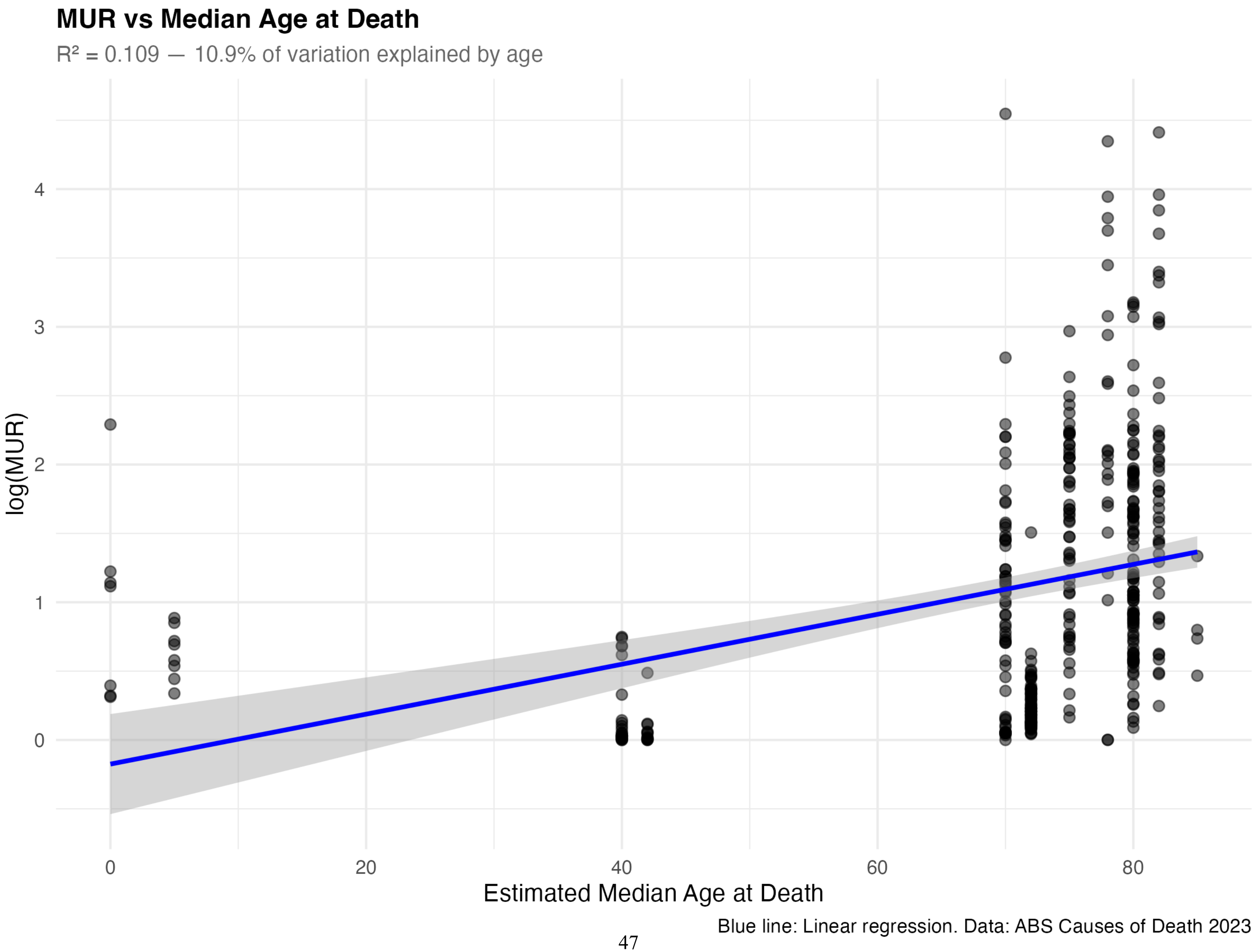

Supplementary Figure S14

**Supplementary Figure S14.** Regression of log(ratio) on estimated median age at death. The fitted line shows the modest positive relationship: older conditions tend to have higher ratios, but the effect is weak ($R^2 = 0.109$). Conditions with large positive residuals have high ratios after accounting for age.

**Supplementary Figure S15. CDC WONDER age-standardisation validation**

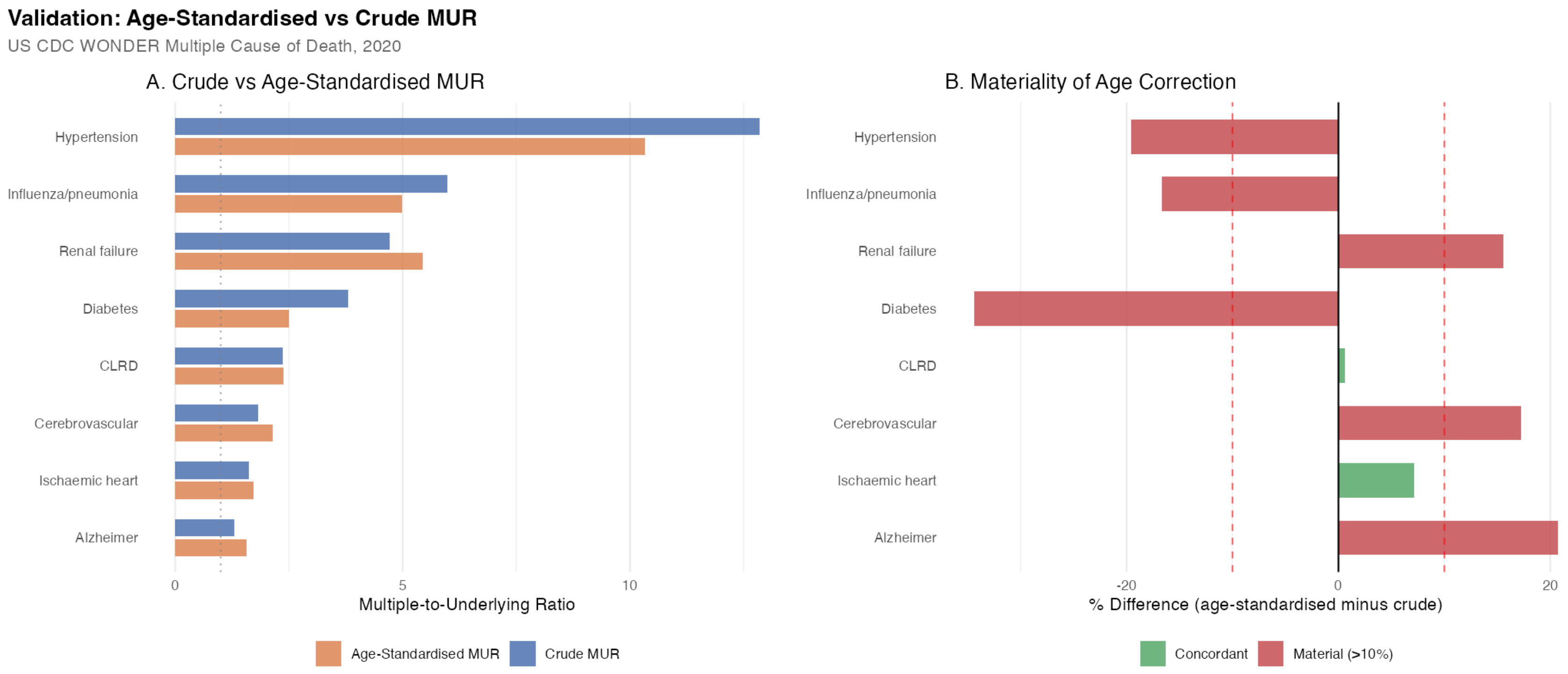

Supplementary Figure S15

**Supplementary Figure S15.** Comparison of crude and age-standardised ratios for eight cause groups using US CDC WONDER data (2020). Bars show crude ratio (dark) and directly age-standardised ratio (light). Conditions where age standardisation substantially changes the ratio are marked with divergence percentages. Only chronic lower respiratory diseases (+0.6%) and ischaemic heart disease (+7.1%) showed concordant estimates; the remaining 6 causes showed material divergence (>10%), indicating that the crude ratio may overestimate or underestimate hidden burden depending on the age profile of decedents.

### Supplementary Data Files

The following data files are available in the online repository (https://github.com/hayden-farquhar/aus-mortality-hidden-burden):

- `mur_ranking_full.csv` (663 rows): Complete ratio ranking for 663 conditions. Key columns: rank, icd_code, condition_name, ratio_persons, ratio_male, ratio_female, sex_ratio.
- `sex_stratified_hidden_burden.csv` (~1,400 rows): Sex-stratified ratio for all conditions. Key columns: icd_code, ratio_male, ratio_female, ratio_diff.
- `holm_bonferroni_results.csv` (8 rows): Multiple testing correction results. Key columns: test_id, p_uncorrected, p_holm_primary, p_holm_all.
- `deaths_underlying_vs_multiple.csv` (~2,000 rows): Core ratio computation table. Key columns: icd_code, underlying_persons, multiple_persons, ratio_persons.
- `state_variation_chapters.csv` (~20 rows): Geographic CV by ICD-10 chapter. Key columns: cause_name, cv, min_state, max_state.
- `deaths_by_num_causes.csv` (~2,000 rows): Death certificate complexity data. Key columns: cause_icd10, reported_alone, with_1_other, etc.
- `multimorbidity_complexity.csv` (~300 rows): Multimorbidity analysis. Key columns: cause_name, pct_alone, pct_with_3plus, avg_co_causes.
- `age_adjusted_mur_ranking.csv` (~200 rows): Age-adjustment sensitivity analysis. Key columns: icd_code, ratio_raw, ratio_adjusted, rank_change.
- `cdc_mur_validation_results.csv` (8 rows): CDC WONDER age-standardisation validation. Key columns: cause_short, crude_ratio, age_std_ratio, pct_difference, material.
